# Mendelianization: Concentrating Polygenic Signal into a Single Causal Locus

**DOI:** 10.1101/2025.10.31.25339237

**Authors:** Eric V. Strobl

## Abstract

Complex disorders such as depression and alcohol use involve numerous genetic variants, and implicated loci continue to grow with sample size. This proliferation hampers interpretability, as the mechanisms by which so many variants jointly contribute to pathophysiology remain unclear. In contrast, classical Mendelian diseases arise from a single causal locus and are easier to interpret. We thus introduce *Mendelianization* – an algorithm distinct from Mendelian randomization – that learns weighted combinations of outcomes so that each aggregated phenotype concentrates association at one locus. We prove that this locus is causal under four structural assumptions natural to genetic data. The method handles partial sample overlap, provides calibrated hypothesis tests, maps coefficients to interpretable scales, and quantifies the degree of Mendelianism using summary *z*-statistics alone. In experiments, Mendelianization enhances statistical power to detect Mendelian symptom profiles even in heterogeneous disorders like major depression, generalized anxiety, and alcohol use disorder. An R implementation is available at github.com/ericstrobl/Mendelianization.

## Introduction

Complex diseases arise from diverse genetic and environmental risk factors. As a result, genome-wide association studies (GWAS) have identified hundreds of associated variants, with larger sample sizes only expanding the catalog.^36^ However, the accumulation of associations often complicates rather than clarifies disease mechanism. Even after fine-mapping prioritizes putative causal variants within loci,^26^ the genetic architecture remains dispersed across many regions and resists reductionist interpretation.^30, 16^ A major culprit is phenotypic heterogeneity, where complex disorders like depression manifest as different symptom constellations (e.g., low mood, anhedonia, fatigue, insomnia). Conventional GWAS collapse this diversity into a single diagnosis or composite, broadening the phenotype and inflating the number of hits without sharper insight.^8, 36^

Existing multi-trait methods do not reduce this proliferation of associated loci. Structure-learning methods (e.g., Genomic SEM,^9^ PDR,^2^ SHAHER,^32^ FactorGo^38^) learn latent outcomes that preserve trait-variant covariance, typically yielding even more polygenic, diffuse signals rather than locus-specific effects. By contrast, power-seeking methods (e.g., HIPO,^20^ metaCCA,^4^ fastASSET^21^) combine traits to maximize a detection criterion such as a non-centrality statistic, identifying still even more variants rather than clarifying mechanism; their nominal *p*-values also tend to be anti-conservative because the outcome is learned on the same data used for testing. Meanwhile, causal inference methods either refine signals within loci (multi-trait fine-mapping^1, 39^) or estimate exposure-outcome relations (multi-response MR^41^ and DAG methods^40^), rather than trying to identify variant-to-outcome causal effects genome-wide.

We thus advance an alternative approach that leverages multiple outcomes to simplify – not expand – the set of associated variants by learning weighted outcome combinations that approximate Mendelian-like traits whose genetic associations concentrate in a single causal locus (Figure 1). This framing treats a heterogeneous disorder as an aggregate of partially distinct symptom dimensions, allowing different loci to anchor different symptom profiles rather than forcing all loci to act through the same diagnosis or total score. We specifically make the following **contributions**:

**Fig. 1.**
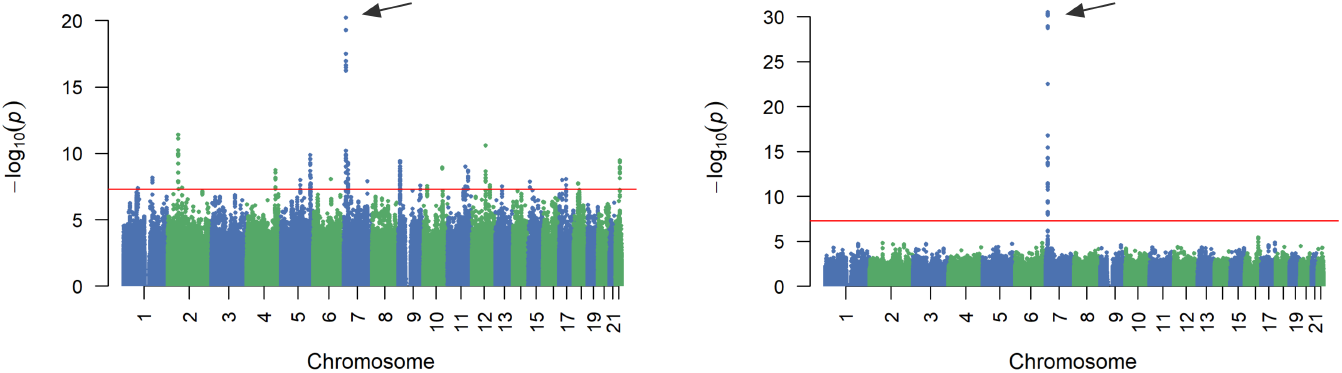
Main idea. Standard outcomes – such as diagnosis or total symptom severity – associate with many loci and obscure underlying mechanisms (left). Mendelianization instead learns an outcome whose association converges on a single locus, yielding a solitary Manhattan plot tower (right). The algorithm thus treats heterogeneous disorders as mixtures of partially distinct symptom dimensions and asks which symptom profile, if any, is most specifically associated with each locus. In practice, **multiple** such outcomes are inferred, one for each lead variant. The red line marks genome-wide significance (5 *×* 10^−8^).

1. We introduce *Mendelianization*, a new framework – distinct from Mendelian randomization – that constructs composite outcomes which concentrate associations at a single locus via a substantially modified canonical correlation analysis (CCA).
2. We prove that this locus contains at least one causal variant under just four structural assumptions.
3. We develop a corresponding fast hypothesis test that remains valid after outcome learning.
4. We operationalize Mendelianization at scale by addressing missing data, enhancing coefficient interpretability, ensuring accurate inference, and introducing a continuous *Score of Mendelianism* to quantify achieved Mendelian properties.
5. We demonstrate that Mendelianization uncovers interpretable Mendelian-like traits in complex disorders using symptom-level summary *z*-statistics.^11^

Together, these advances provide a reductionist framework for identifying Mendelian-like symptom profiles dominated by a single causal locus.

## Methods

### One-Dimensional Canonical Correlation Analysis

One-dimensional CCA will serve as the underlying engine of Mendelianization. We thus consider summary statistics comprised of the partial correlation matrix *r* ∈ ℝ^*q*×*m*^ between *q* variants *V* and *m* outcomes *Y*. We assume that all covariates like biological sex, age, and ancestry have already been partialed out from *V* and *Y*. We focus on population-level properties for ease of presentation and address sample-level issues later. We thus have *r*_*jk*_ = cor(*V*_*j*_, *Y*_*k*_). Let *r*_*j*_ correspond to the row of *r* associated with the variant *V*_*j*_.

For each *V*_*j*_, we construct a composite outcome *Y α*_*j*_ by one-dimensional CCA, so that the correlation between *V*_*j*_ and *Y α*_*j*_ is as large as possible:

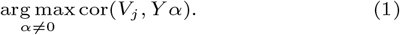

We will also show that this procedure implicitly reduces the correlations between loci not containing *V*_*j*_ and *Y α*_*j*_, so that *Y α*_*j*_ corresponds to a Mendelian-like trait.

Without loss of generality, we assume that each variant and outcome has already been standardized to mean zero and unit variance. We can then simplify the above objective to:

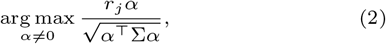

where Σ corresponds to the positive definite correlation matrix of *Y*. Notice that the numerator maximizes alignment with *V*_*j*_, while the denominator minimizes the scale of *α* according to the geometry of Σ.

The solution to Expression (2) is the familiar CCA weight vector that whitens the outcomes and then aligns with *r*_*j*_. In particular, up to an arbitrary scale factor, the optimal weights satisfy

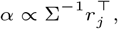

so we set 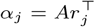 with *A* = Σ^−1^ from here on. We provide a detailed derivation of this solution in Supplementary Materials A.

### Causal Analysis

We next ask when the CCA-derived composite *Y α*_*j*_ admits a causal interpretation. In particular, we show that, under a set of structural assumptions motivated by genetic data, the optimization problem in Expression (1) produces a composite outcome whose association pattern is concentrated within a single linkage disequilibrium (LD) block, and that this block must contain at least one causal variant for *Y α*_*j*_. We refer to this property as *Mendelianization*.

#### Structure of Latent Confounding

Even after adjusting for observed covariates such as age, sex, and ancestry, genetic association studies typically retain residual confounding from processes such as batch effects, residual population structure, and dynastic effects.^36^ These processes tend to act on many phenotypes at once and therefore induce shared structure across outcomes.

We formalize this by assuming:

##### Assumption 1.

*(Low-dimensional latent confounding) For sufficiently large m, the remaining confounding effects lie in a subspace U*^*m*^ ⊂ ℝ^*m*^, *and there exists an integer constant r*_0_ ≤ 0 *such that* dim(*U*) ≥ *r*_0_.

**Remark**. Assumption 1 states that *V* and *Y* are influenced by only a small number of latent confounders, or by many confounders whose effects span a low-dimensional subspace. This matches empirical experience in large biobanks, where a handful of unmeasured processes generate broad, shared patterns across many traits. For example, an environmental exposure may correlate with allele frequencies at some loci and affect several symptoms. This setting remains compatible with Assumption 1 when the induced symptom-level confounding pattern is low-dimensional. However, Assumption 1 would be violated if there were many environmental confounding effects whose symptom patterns were high-dimensional and not described by a shared low-rank structure.

Assumption 1 implies that each vector of variant-outcome correlations 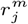 can be written as

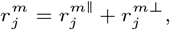

where 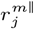 lies in the confounding subspace *U*^*m*^ and 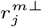 lies in its orthogonal complement (with respect to *A*^*m*^ = (Σ^*m*^)^−1^). Informally, 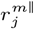 captures residual confounding, while 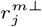 captures the causal component of the association between *V*_*j*_ and the outcomes. We use this decomposition to study how Mendelianization behaves with and without latent confounding.

#### Without Latent Confounding

We first consider the idealized case where latent confounding is absent, so 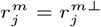 for all *j*. Let 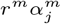 denote the vector of associations between each variant and the composite outcome defined by 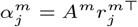. For each variant *V*_*k*_,

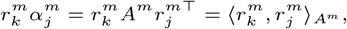

where 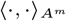 denotes the inner product induced by *A*^*m*^. When *k* = *j*, this quantity measures how strongly *V*_*j*_ associates with its own learned outcome. When *k j*, the same expression measures the alignment between the causal pattern of *V*_*j*_ and that of *V*_*k*_ across the *m* outcomes.

To obtain a single-locus “tower” in the Manhattan plot of 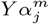, we require that this alignment vanish for variants outside the LD block around *V*_*j*_. Let S_*j*_ denote the set of variants in LD with *V*_*j*_ (the locus or LD block containing *V*_*j*_). We assume:

##### Assumption 2.

*(Outcome diversity) We have*

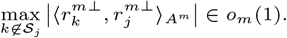

**Remark**. In the absence of confounding, 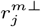 represents the causal pattern of effects of *V*_*j*_ on the outcomes. Assumption 2 concerns these patterns in the *A*^*m*^ = (Σ^*m*^)^−1^-whitened outcome space, not in the original correlated outcome coordinates. Thus, as we add more non-redundant outcomes, the whitened causal patterns from different loci become increasingly orthogonal. Mixed positive and negative effects from variants outside S_*j*_ tend to cancel across outcomes, driving their contribution to 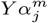 toward zero. By contrast, variants in S_*j*_ share the same underlying causal mechanism for 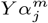 and therefore retain strong alignment. This is plausible in high-dimensional phenotyping because adding more granular traits (e.g., subphenotypes or symptom items) typically increases the heterogeneity of causal effects across loci. In sum, the vector 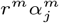 develops a sharp tower within S_*j*_ as *m* grows, while associations at other loci approach zero.

#### With Latent Confounding

In real data, latent confounding is present and can also create tall peaks in Manhattan plots. For example, the composite 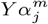 might load on a pattern driven purely by confounding, such that

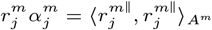

even if no variant in S_*j*_ is truly causal for 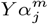. To rule out such purely confounded towers, we impose additional assumptions on the strength of both causal and confounded components.

We first assume that the relevant signals do not vanish as we add more outcomes:

##### Assumption 3.

*(Signal floors) For sufficiently large m, if l* ∈ S_*j*_ *causally influences* 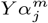, *then there exists a constant β*_c_ *>* 0 *such that*

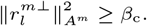

*Moreover, there exists a constant β*_on_ *>* 0 *such that*

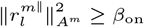

*for every l* ∈ S_*j*_ *that does not cause* 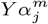.

**Remark**. The first part of Assumption 3 requires that if S_*j*_ contains a causal variant for *Y α*_*j*_, its causal signal remains non-negligible even as we increase the number of outcomes. This is a minimal requirement for any causal effect to be detectable. The second part states that non-causal variants in the same LD block share a non-vanishing confounded component, because LD makes them inherit similar exposure to the same local confounders (such as local ancestry or technical artifacts). This is consistent with how confounding typically behaves within LD regions in GWAS.

We also formalize how low-dimensional confounding propagates across the genome:

##### Assumption 4.

*(Isolation-spillover) If, for all sufficiently large m, some l*_*m*_ ∈ S_*j*_ *causally influences* 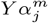, *then*

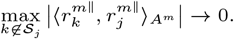

*Conversely, for every sufficiently large m such that no l* ∈ S_*j*_ *causes* 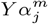, *there exists a variant k*_*m*_∉ S_*j*_ *such that*

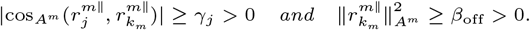

**Remark**. The first clause states that when S_*j*_ contains a causal variant for *Y α*_*j*_, the associated confounding pattern does not align strongly with distant loci, so any remaining confounding is effectively local to the LD block. The second clause describes the opposite scenario: if the tower at S_*j*_ is driven *purely* by low-dimensional confounding, then at least one other locus must share the same confounding pattern with non-negligible strength, leading to detectable “spillover” of confounded signal elsewhere in the genome. This reflects the empirical behavior of broad confounding sources – such as uncorrected population structure, ascertainment, site, batch, or subgroup-level environmental differences – which typically induce shared association patterns across many loci rather than a single isolated peak. Thus, if a subgroup of individuals shares both an environmental exposure and elevated frequency of a variant at a particular locus, Assumption 4 does not require this structure to be absent; it requires that such noncausal subgroup-associated signal also spill over to other loci carrying similar subgroup-associated patterns, rather than masquerading as a clean isolated single-locus tower.

#### Asymptotic and Finite Mendelianism

Under Assumptions 1–4, the CCA estimator enjoys a strong causal localization property:

##### Theorem 1.

*(Asymptotic Mendelianism) Consider Assumptions 1–4 and suppose* Σ^*m*^ *is positive definite for sufficiently large m. The following statements are equivalent:*

i. *There exists a constant c*_*j*_ *>* 0 *such that* 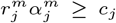 *for sufficiently large m, and* 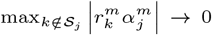, *so Mendelianism is achieved asymptotically*.
ii. *For all sufficiently large m, there exists at least one variant l*_*m*_ ∈ S_*j*_ *that causes* 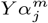.

**Remark**. We provide all proofs in the Supplementary Materials. Theorem 1 establishes both directions: the composite outcome 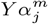 becomes asymptotically Mendelian-like at the LD region S_*j*_ if and only if, for all sufficiently large *m*, this region contains at least one causal variant for 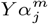. Intuitively, under the four structural assumptions (low-dimensional latent confounding, outcome diversity, signal floors, and isolation-spillover), maximizing the correlation between *V*_*j*_ and *Y α* forces the resulting composite outcome to localize its causal signal to a single LD block. This contrasts with common practice, which fixes a single outcome or factor and tests all variants, often yielding many significant loci scattered across the genome (Figure 1, left). By comparison, the estimator 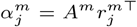 learns a variant-specific composite outcome for each *V*_*j*_, and Theorem 1 shows that the learned outcome 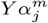 has asymptotically vanishing associations outside one LD region and that this region contains at least one causal variant for the learned outcome (Figure 1, right).

Beyond this localization guarantee, Theorem 1 also changes how we interpret CCA loadings. For each variant *V*_*j*_, we estimate the canonical coefficients 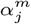 and form the composite 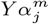. **We then hold this composite fixed and evaluate association genome-wide** – not just at *V*_*j*_ itself. Each 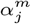 thus defines a Mendelianized trait that we scan across all variants. We interpret 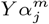 as a Mendelian-like trait when the corresponding Manhattan plot shows a single tower confined to an LD region containing *V*_*j*_.

Finally, we recover a stronger version when a finite outcome set is already sufficiently rich:

##### Proposition 1.

*(Perfect Mendelianism at finite m) Consider Assumptions 5–8 (Supplementary Materials B) with a prespecified m and suppose* Σ^*m*^ *is positive definite. The following statements are equivalent:*

i. *We have* 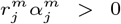 *and* 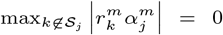, *so Mendelianism is perfectly achieved for S*_*j*_ *at m*.
ii. *At least one variant l* ∈ S_*j*_ *causes* 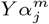.

**Remark**. Proposition 1 shows that, under stronger assumptions, Mendelianization can in principle recover a classical single-locus architecture for the learned outcome at a fixed number of outcomes *m*. We emphasize Theorem 1 because it uses weaker, more realistic assumptions and allows Mendelianism to emerge asymptotically as we enrich the outcome set. This distinction is important for complex, polygenic diseases, where perfect single-locus resolution may not be achievable at any fixed *m*.

### Fast Hypothesis Testing

We now test the null of no dependence between *V*_*j*_ and *Y*, even though we learn *α*_*j*_ from the same data. We henceforth drop the superscript *m* to simplify notation. The null and alternative hypotheses are:

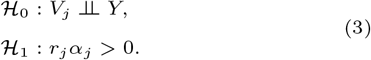

A naive Pearson test would be anti-conservative, because it treats *α*_*j*_ as fixed even though we estimated it using the observed associations. To obtain a valid test, we construct a statistic whose null distribution accounts for the fact that *α*_*j*_ is data-driven.

Under ℋ_0_, with i.i.d. samples and finite fourth moments,

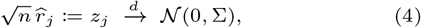

where *n* denotes sample size^1^. The canonical coefficients are proportional to 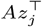, which motivates the quadratic form 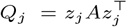.

To understand the null distribution of *Q*_*j*_, write the Cholesky decomposition Σ = *LL*^⊤^ and let *u* ~ *N*(0, *I*_*m*_) so that *z*_*j*_ = *u*^⊤^*L*^⊤^. Substituting into *Q*_*j*_ gives

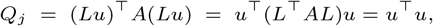

so that 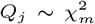 under *H*_0_. We then compute the right-tail probability of *Q*_*j*_ under the 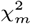 distribution to obtain the *p*-value.

The above *χ*^2^ result is asymptotic, and its finite-sample accuracy depends on how well we approximate Σ. In the next section, we show that a modified version of Σ can be consistently estimated using millions of variants under realistic LD and sparsity conditions, so that large *n* and large *q* asymptotics work decisively in our favor.

### Operationalizing Mendelianization at Scale

Implementing the above hypothesis test requires access to Σ. However, biobanks rarely report Σ in their summary statistics, and the outcomes needed to estimate Σ are typically measured on partially overlapping sets of individuals. Scaling Mendelianization also raises two additional challenges beyond test calibration: redundant traits can obscure the interpretation of raw canonical coefficients, and the available traits may not be rich enough to approach near-perfect Mendelianism. We therefore (a) replace Σ with a quantity that is estimable from summary *z*-statistics under partial overlap, (b) show how to estimate it at scale despite LD, (c) rescale coefficients for interpretability, and (d) introduce a metric to quantify how close a locus comes to Mendelian behavior.

#### From Σ to Γ Under Partial Overlap

If all traits are fully observed and *V* ⫫ *Y*, then 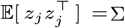 with i.i.d. sampling, and we can consistently estimate Σ using (approximately) independent variants. In practice, samples overlap only partially across traits, so *z*-statistics and correlations are computed from unequal sample sizes.

Let *M*_*a,i*_ ∈ {0, 1} indicate whether *Y*_*a,i*_ is observed, and define *S*_*a*_ = {*i* : *M*_*a,i*_ = 1}, *n*_*a*_ = |*S*_*a*_|, and *n*_*ab*_ = |*S*_*a*_ ∩ *S*_*b*_|. We assume that *V* contains no missing values, and that each *Y*_*a*_ has been centered so that *E*(*Y*_*a*_|*M*) = 0 for the realized missingness pattern *M*. Let

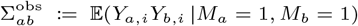

denote the (*a, b*) pairwise observed second-moment entry, conditional on both *Y*_*a*_ and *Y*_*b*_ being observed. Then:

##### Proposition 2.

*Assume i*.*i*.*d. samples across individuals, V* ⫫ *Y, and V* ⫫ *M* | *Y*. *Then* 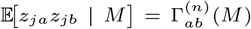,

*where*

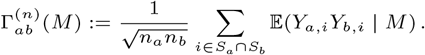

*Separately, without requiring V* ⫫ *Y or V* ⫫ *M* | *Y, under i*.*i*.*d. missingness, with p*_*a*_ = *P* (*M*_*a*_ = 1) *>* 0, *p*_*b*_ = *P* (*M*_*b*_ = 1) *>* 0, *and p*_*ab*_ = *P* (*M*_*a*_ = 1, *M*_*b*_ = 1) *>* 0,

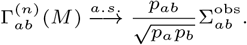

**Remark**. The assumption *V* ⫫ *M* | *Y* is reasonable in biobank settings since missingness in *Y* is largely driven by the presence or severity of symptoms themselves (as well as factors such as sex, age, and ancestry, which we already adjusted for). Proposition 2 also motivates replacing Σ with the matrix Γ in Expression (2), where Γ denotes the large-sample limit of the realized conditional covariance matrix Γ^(*n*)^(*M*), with entries

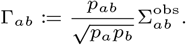

Replacing Σ with Γ has a convenient interpretation. Define modified outcomes

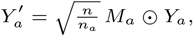

which set missing entries of *Y*_*a*_ to zero and rescale by the square root of the observed sample proportion. Then *E*(*Y*′|*M*) = 0 for the realized *M*, and hence 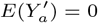. Moreover:

##### Proposition 3.

*Assume i*.*i*.*d. samples across individuals, p*_*a*_ *>* 0, *p*_*b*_ *>* 0, *p*_*ab*_ *>* 0, *and the corresponding conditional product moments are uniformly integrable. Then*

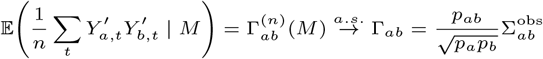

*and*

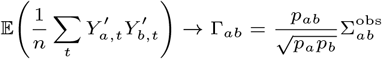

*as n* → ∞.

**Remark**. 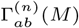 is therefore the *conditional* covariance of *Y*′ for the realized missingness pattern *M*, and it converges to Γ_*ab*_, the *unconditional* covariance of *Y*′, as sample size grows. As a result, we can maximize the (asymptotically) unconditional correlation in Expression (2) by working with *Y*′ instead of *Y* :

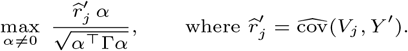

Importantly, this reparameterization targets Γ directly and therefore **does not require missingness-at-random (MAR)**^13^ for Σ.

Theorem 1 then holds for *Y*′ at the population level with Σ replaced by Γ. The scaling factors 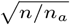 used to define *Y*′ do not affect the optimization, since:

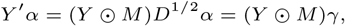

where *D* = diag(*n/n*_1_, …, *n/n*_*m*_) and *γ* = *D*^1/2^*α*. Hence:

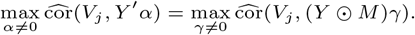

Even better, we never need explicit access to *Y*′ because the sample covariance simplifies to:

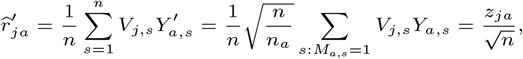

which implies:

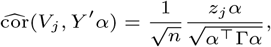

except that we replace 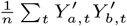 in the denominator with Γ, the large-sample limit of its conditional counterpart Γ^(*n*)^(*M*), by Proposition 3. Thus, we only need the *z*-statistics *z*_*j*_ and an estimate of Γ.

This reasoning motivates us to maximize:

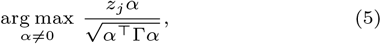

which mirrors Expression (2) with Σ replaced by Γ. We henceforth take 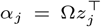 as our solution, where Ω = Γ^−1^. Importantly, this objective still maximizes the association between *V*_*j*_ and the learned composite *Y α*; it does not directly optimize genome-wide localization.

#### Estimating Γ with LD

Although Γ is natural to target, empirical estimation is nontrivial. In principle, Proposition 2 shows that genome-wide *z*-statistics identify the realized conditional covariance Γ^(*n*)^(*M*) under the global null *V* ⫫ *Y*, whose large-sample limit is Γ. In practice, two main obstacles remain: LD induces dependence between nearby variants, and a subset of variants truly associate with *Y*. Fortunately, we can handle both these issues with the following result:

##### Theorem 2.

*For fixed outcomes a and b, consider the estimator* 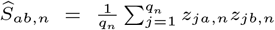, *where q*_*n*_ → ∞ *as n* → ∞. *Let G*_*n*_ *index the variants that are independent of Y, and let B*_*n*_ *index the variants that are dependent on Y*. *Define X*_*j,n*_ = *z*_*ja,n*_*z*_*jb,n*_. *Assume the conditions of the second part of Proposition 2 hold and further assume:*

1.*(Asymptotic null centering) There exists a sequence η*_*n*_ → 0 *such that* 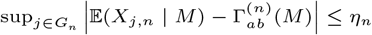

2.*(Vanishing aggregate contamination) We have*

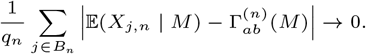

3.*(Local dependence) Conditional on M, the variables* 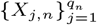 *admit an undirected dependency graph with maximum degree D, and* 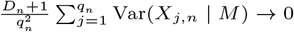.

*Then* 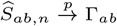.

**Remark**. The key condition is the local LD assumption. In practice, investigators routinely treat LD as local by bounding each variant’s LD neighborhood, for example within 1 Mb, which makes the dependency degree *D* finite. Under this condition, the variance of Ŝ_*ab*_ goes to zero as *q* → ∞. Moreover, the vanishing contamination assumption reflects the sparse genetic architecture of most traits: after standard quality control, only a tiny fraction of variants are typically associated with any given outcome, so the non-null set *B*_*q*_ grows more slowly than *q*. This matches empirical experience in large biobanks, where the vast majority of variants behave as null.

Taken together, these conditions imply a weak law of large numbers despite LD. In practice, with millions of variants and large sample sizes, we can estimate each entry of Γ to negligible error. We therefore treat Γ as known in subsequent derivations. Consequently, under H_0_, 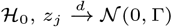, and we replace Σ with Γ and *A* with Ω = Γ^−1^ so that 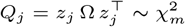 in the test of Expression (3). We will later show that this test works very well in real data.

#### Interpretable Coefficients

We can now solve Expression (5) using an estimate of Γ. However, the raw coefficients correspond to:

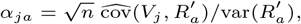

where 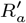 is the residual of 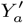 after regressing it onto 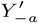, so they depend on the residual variance 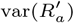, which varies across outcomes and complicates comparisons.

To address this, we report rescaled coefficients 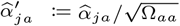 for interpretability:

##### Proposition 4.

*We have* 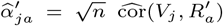 *for every Y*_*a*_ ∈ *Y*.

Thus, 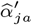 equals 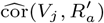 up to the common factor 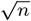. The rescaled canonical coefficients are unitless, directly comparable across outcomes for a given variant *V*_*j*_, and their signs match the allelic direction of effect on the unique component of 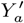 (the residual 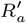 after adjusting for the other traits). In practice, 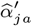 can be viewed as a standardized effect of *V*_*j*_ on the part of trait *a* that is not explained by the remaining traits in *Y*.

The unique-component interpretation reduces redundancy among correlated outcomes and makes the coefficient pattern easier to interpret. For example, if one symptom partly reflects both anxiety and low mood, whereas another symptom mainly reflects low mood, then the residualized component of the first symptom more specifically captures the anxiety-related variation not shared with the low-mood symptom.

The signs of the coefficients are interpreted relative to the allele dosage used to compute the input *z*-statistics. Positive entries of 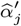 indicate unique symptom components associated with higher effect-allele dosage, whereas negative entries indicate unique symptom components negatively associated with higher effect-allele dosage. The vector 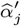 should therefore be interpreted jointly as an allele-oriented, locus-anchored profile of associations with unique symptom components. We provide concrete examples of coefficient interpretation in the Results.

#### Quantifying the Degree of Mendelianism

Near-perfect Mendelianism rarely occurs when the outcome set is small (for example, *m* ≤ 15). Although Theorem 1 provides a yes/no criterion in the limit, applied analyses benefit from a continuous measure that captures how close *Y α*_*j*_ approaches Mendelian-like behavior.

We therefore fix the coefficients *α*_*j*_ learned at the lead variant *V*_*j*_ and, for any variant *V*_*k*_ (including *k* = *j*), compute

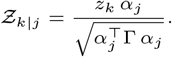

Under *H*_0,*k*_ : *r*_*k*_*α*_*j*_ = 0, we have 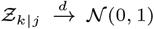 with i.i.d. samples and finite fourth moments, yielding the two-sided *p*-value *p*_*k*|*j*_ = 2Φ −|Z_*k*|*j*_|^2^. We compute Z_*k*|*j*_ and *p*_*k*|*j*_ for all *V*_*k*_ ∈ *V* (i.e., genome-wide).

If we reject H_0_ in Expression (3) for *V*_*j*_, we summarize how concentrated the signal is at the *j*-locus with the **Score of Mendelianism (SoM)**:

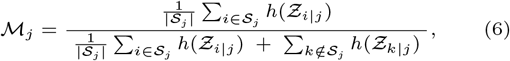

where *h*(*Ƶ*) = (|*Ƶ*| − *z*_*α*_)_+_, and *z*_*α*_ is the *z*-score corresponding to a two-sided *p*-value (default *α* = 5 × 10^−8^). We construct S_*j*_ using proximity-based clustering around *V*_*j*_ (single-linkage with 500 kb gap, padded by ±500 kb), though alternatives such as LD-based clustering relative to *V*_*j*_ are also possible.

**Remark**. The SoM lies in [0, 1] and can be interpreted as the fraction of “excess” Mendelianized signal (above the genome-wide threshold *z*_*α*_) that lies within the LD region of interest. A value of *M*_*j*_ = 1 indicates complete isolation of the signal to *S*_*j*_ and therefore provides the strict criterion for a perfectly localized Mendelian-like trait. Values below one indicate approximate Mendelianism: higher values correspond to greater concentration of signal at *S*_*j*_, even when some off-target signal remains. Conversely, *M*_*j*_ approaches 0 when the signal lies entirely outside *S*_*j*_ or is highly polygenic. We therefore use *M*_*j*_ primarily as a continuous descriptive and ranking measure among loci that are already genome-wide significant, rather than imposing an arbitrary universal cutoff below one. We average within *S*_*j*_ to avoid favoring larger loci and *do not* average outside *S*_*j*_, so that even small off-target regions remain visible rather than being diluted across millions of variants.

In practice, we use *M*_*j*_ only as a ranking metric among loci that are already genome-wide significant. Formal association testing is performed using the 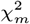 statistic, which accounts for learning *α*_*j*_ when testing whether *V*_*j*_ is associated with the multivariate outcome. Thus, outcome learning alone should not systematically convert noise or overfitting into significant composite associations, and isolated chance findings are further limited by the stringent genome-wide significance threshold.

The SoM and corresponding Manhattan plots only summarize the fitted genome-wide localization pattern after *α*_*j*_ has been fixed and applied to all variants. They are not assigned calibrated *p*-values and are not used to control Type I error for localization. Replication or sample splitting is therefore not required for validity of the association test, but can be used to assess whether the observed localization pattern is stable beyond the discovery dataset.

#### Summary

Mendelianization corresponds to one-dimensional CCA, modified in five ways: (1) it uses *z*-statistics in place of raw correlation coefficients, (2) it overcomes missingness by normalizing with Γ estimated over variants rather than with Σ estimated over outcomes, (3) it replaces Wilks’ Λ with a 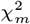 test for significance, (4) it rescales canonical coefficients by the square roots of the diagonals of Ω for interpretability, and (5) it applies each significant composite outcome genome-wide by Theorem 1. Pseudocode and a formal time complexity analysis (Supplementary Materials C) show that Mendelianization scales cubically with the number of outcome variables but only linearly with the number of variants. This remains computationally practical in modern biobanks, where the number of outcomes rarely exceeds 100, whereas the number of variants often surpasses one million.

## Results

### Comparators

We compared Mendelianization against:

- **Meta-analytic Canonical Correlation Analysis (metaCCA)**: performs CCA on summary statistics using Wilks’ Λ, which assumes perfectly overlapping and equal sample sizes per trait. metaCCA also estimates Σ from linear regression *β* coefficients, so it cannot fully combine linear and logistic regression models whose coefficients differ in scale and null variances. Finally, metaCCA lacks causal guarantees, and its scale-dependent canonical coefficients are not directly comparable across traits.
- **Heritability Informed Power Optimization (HIPO)**: learns multiple sets of outcome weights that increase statistical power by maximizing a standardized non-centrality parameter across – rather than within – loci. We compare against the most powerful, first set of HIPO weights.
- **Fast Association analysis based on SubSETs (fastASSET)**: a multi-trait analysis method that first pre-screens traits with suggestive associations and adjusts for this selection. It then searches over all (two-sided) subsets of traits – allowing opposite effect directions – and combines evidence into a single, multiple testing adjusted omnibus *p*-value per variant.

These methods were not designed to optimize single-locus concentration, but they are the closest available comparators because each combines information across multiple traits to improve genetic association detection. Mendelianization can also be viewed as a power-maximizing method, since it learns the outcome direction most strongly associated with each variant. Its additional contribution is that, under the stated assumptions, this variant-specific association maximization yields a learned outcome whose signal concentrates in a single LD region containing at least one causal variant. We finally also compared Mendelianization against **three ablated variants**: (i) using variant-specific outcomes (i.e., Z_*i*|*j*_ = Z_*i*|*i*_) as opposed to fixed outcomes applied genome-wide, (ii) estimating Γ using *β* coefficients instead of *z*-statistics, and (iii) a combination of both (i) and (ii).

### Metrics

Our primary evaluation metric was the **Score of Mendelianism (SoM)**, where higher values indicate a greater ability to isolate a single locus. The metric provides a fair basis for comparison because no algorithm, including Mendelianization, optimizes SoM. Given that Mendelianization and metaCCA have different coefficient interpretations, we computed SoM with variant-specific outcomes for metaCCA, and with fixed outcomes applied genome-wide for Mendelianization (the outcome learned at the lead *V*_*j*_, applied to all variants). This ensures each method is evaluated in line with its intended use.

Beyond detecting a Mendelian-like pattern, rigorous applications also require valid, efficient inference. As a result, we evaluated calibration using the **signed Kolmogorov– Smirnov (KS) distance** between the observed *p*-values above 0.05 and the Uniform[0.05, 1] distribution. Well-calibrated methods should yield approximately uniform *p*-values in this range, because most variants are expected to be null or only weakly associated with the learned outcome. This metric is useful because it can be computed in both synthetic and real-data analyses, unlike conventional Type I error evaluation, which requires a known global null. To complement this real-data-compatible calibration metric, we also performed formal global-null simulations and evaluated the standard KS distance against the full Uniform[0, 1] distribution.

For methods with acceptable calibration, defined as positive anti-conservative KS distances below 0.01, we next assessed a **statistical proxy power**: the proportion of variants with *p <* 10^−4^. This sub-genome-wide threshold summarizes low-*p*-value enrichment among calibrated methods. Although this quantity is not a formal power estimate in real data, where the true non-null variants are unknown, it again provides a practical measure of signal yield that can be computed in both synthetic and real-data analyses. In the synthetic experiments, where the causal variants are known, we additionally evaluated **standard empirical power** as the true positive rate at the genome-wide threshold *p <* 5 × 10^−8^.

We further assessed **composite-score stability** in simulations by generating two independent datasets under the same causal architecture and comparing the recovered locus-specific composite directions across datasets. For each true causal variant, we computed the Γ-weighted cosine similarity between the two independently learned coefficient vectors,

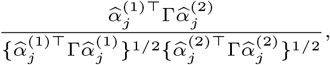

which corresponds to the correlation between the two learned composite scores under the outcome covariance structure. Thus, values near one indicate recovery of the same locus-specific composite direction, rather than exact equality of the raw coefficient weights. This analysis could not be performed in the real-data applications because independent matched summary statistics were not available. Finally, we measured **run-time**.

### Simulations

We first conducted simulations using individual-level autosomal genotype data from European-ancestry participants in the 1000 Genomes Phase 3 dataset.^5^ After regressing out the first five principal components, we partitioned the genome into 1,006 LD blocks using the optimal LD-splitting algorithm^19^ (parameters: *r*^2^ threshold for ignoring pairwise correlations = 0.02, window size = 500 Kb, maximum number of variants per block = 2,000, minimum number of variants per block = 200). We then randomly sampled 100,000 variants and, for each of 40 bootstrap replicates, randomly selected five causal variants *V*_*c*_ from this set. For each replicate, we first generated 50 latent traits according to

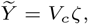

and then constructed 32 observed outcomes,

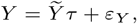

where 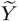 contained 50 variables, each element of *ζ* was drawn from a uniform distribution on [−0.05, −0.01] ∪ [0.01, 0.05], each entry of *τ* was drawn from a standard normal distribution, and *ε*_*Y*_ also followed a standard normal distribution. This construction mimics realistic settings in which observed outcomes are noisy, imperfect measurements that do not map cleanly onto the underlying biology. For each outcome, we randomly drew the sample size from a uniform distribution between 50,000 and 100,000 individuals by bootstrap resampling individuals from 1000 Genomes. Within each LD block, we then performed a second-level bootstrap to disrupt cross-block correlations and to approximate GWAS-scale sampling variability given the relatively small 1000 Genomes sample size. We applied all methods to the resulting 40 simulated datasets for each outcome configuration (2, 4, 8, 16, and 32 outcomes). For the global-null simulations, we used the same genotype sampling, outcome-mixing, sample-size, and blockwise bootstrapping pipeline, but replaced the genotype-driven latent traits 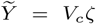 with independent noise traits 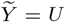, where the entries of *U* were drawn independently from a standard normal distribution. Thus, the observed outcomes retained the same mixing and noise structure as in the main simulations, but contained no genotype-driven signal.

We summarize the simulation results in Figure 2, using primarily violin plots. Mendelianization achieved the highest SoM across all numbers of outcomes (Figure 2 (a)). Consistent with Theorem 1, the SoM steadily increased toward one as the number of outcome variables increased, indicating that the algorithm identified increasingly Mendelian-like traits with more outcomes. In contrast, the other methods produced SoMs near zero, reflecting the fact that we generated the data from five randomly chosen causal variants rather than a single causal variant.

**Fig. 2.**
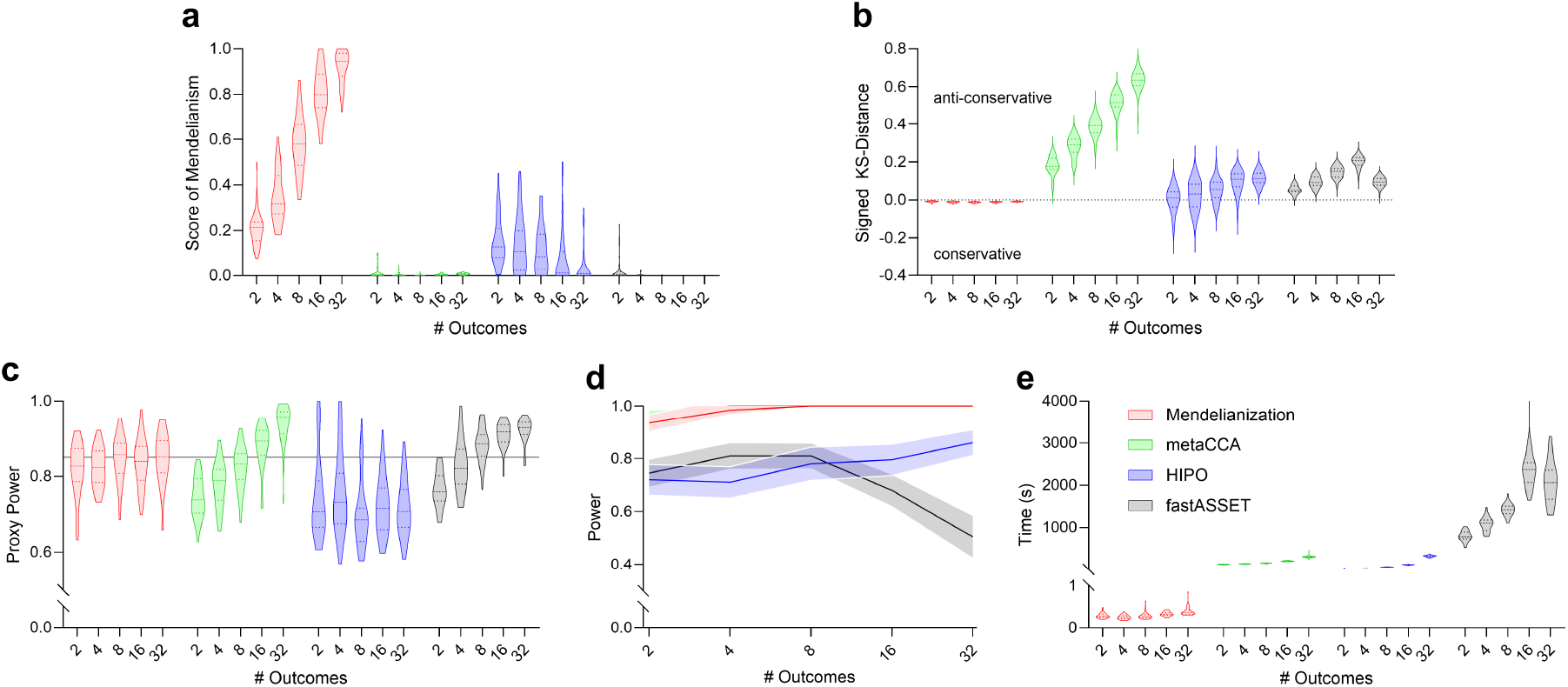
Main results from the synthetic data experiments. All plots share the same legend. (a) Mendelianization achieved the highest SoM across all numbers of outcomes, with SoM increasing as the number of outcomes grew, consistent with Theorem 1. (b) Mendelianization was the only algorithm that produced well-calibrated *p*-values, in agreement with Theorem 2. (c,d) Mendelianization consistently maintained proxy power at approximately 0.85 and achieved near-perfect empirical power at the genome-wide threshold across all outcome dimensions. Panel (d) is displayed as a line plot because the mean true positive rate assumes only a discrete set of values. Error bars denote 95% confidence intervals for the mean. (e) Mendelianization completed in under one second for all runs.

Mendelianization also yielded the most well-calibrated *p*-values for null variants, with calibration close to ideal, consistent with Theorem 2 (Figure 2 (b)). By comparison, most alternative algorithms showed progressively poorer calibration as the number of outcomes increased, indicating a failure to control the type I error rate under outcome learning. Mendelianization also retained good calibration in the null simulations (Supplementary Figure 2 (a)). Across all null simulations, Mendelianization identified no genome-wide significant variants.

Mendelianization was further distinguished as the only method that consistently maintained proxy power at approximately 0.85 across all outcome dimensions (Figure 2 (c)), whereas competing approaches fluctuated substantially, consistent with their less stable *p*-value calibration. Direct ground-truth power at the genome-wide threshold *p <* 5 × 10^−8^ also confirmed that Mendelianization was statistically powerful, increasing from 0.935 at *m* = 2 to 1.000 at *m* ≥ 16 (Figure 2 (d); detailed bar plots in Supplementary Figure 2 (b)).

In terms of numerical and composite-score stability, the condition number of Γ remained below 51 across all outcome dimensions, suggesting that the matrix inversions were numerically stable (Supplementary Figure 2 (c)). Mean composite stability was likewise very high, with Γ-weighted cosine similarities ranging from 0.980 to 1.000, indicating that Mendelianization recovered nearly identical locus-specific composites under independent perturbations (Supplementary Figure 2 (d)). Thus, although the condition number increased with the number of outcomes, empirical composite stability remained essentially unchanged. Finally, Mendelianization completed in under one second for all runs, whereas other algorithms required orders-of-magnitude longer computation times (Figure 2 (e)).

Overall, the results indicate that Mendelianization identifies more Mendelian-like traits as the number of outcomes grows, maintains well-calibrated *p*-values with high power, and remains computationally efficient. Ablation results confirmed the necessity of applying canonical coefficients genome-wide and using *z*-statistics to achieve maximal performance (Supplementary Figure 1). Sensitivity analyses incorporating latent confounding produced qualitatively similar results (Supplementary Materials; Supplementary Figures 3 and 4).

### Depression and Generalized Anxiety

We next evaluated the algorithms using real European ancestry summary statistics from the Pan-UK Biobank.^3, 11^ The outcome variables comprised all 52 non-collinear items in the UK Biobank Mental Health Questionnaire assessing lifetime major depression and generalized anxiety. For all experiments, we restricted analyses to high-quality HapMap3 autosomal, non-MHC, biallelic SNPs with INFO*>*0.9 and MAF*>*1% in UK Biobank and, when available, gnomAD genome/exome. The condition number of Γ was 45.3, suggesting that inversion of Γ was numerically stable for this analysis.

Figure 3 summarizes the results. Mendelianization identified three loci that surpassed the significance threshold of 5 × 10^−8^. These loci also attained perfect SoMs of one, whereas most loci identified by the other algorithms had scores close to zero (Figure 3 (a)). Applying each of the three learned composite *Y α*_*j*_ genome-wide and plotting the *p*-values associated with the *z*-statistics Z_·|*j*_ yielded a single tower per locus, as implied by the SoMs and predicted by Theorem 1 (Figure 3 (b)). The signed interpretable coefficients further delineated three distinct effect-allele-oriented depression/anxiety profiles: (i) role-impairing irritable anxious distress, characterized by higher anxiety-related role impairment, irritability, restlessness, sleep disturbance, and difficulty stopping worry; (ii) fatigue-weight-gain persistent worry, characterized by higher fatigue, persistent worry, concentration difficulty, depression affecting a larger fraction of the day, and depression-related weight gain; and (iii) episodic weight-loss depression, characterized by higher depression-related weight loss, concentration difficulty, and self-treatment, but lower lifetime number of depressive episodes (Supplementary Figure 5). Together, these results show that Mendelianization achieved the greatest spikedness while also recovering interpretable coefficient patterns that separated clinically meaningful dimensions of depression and anxiety.

**Fig. 3.**
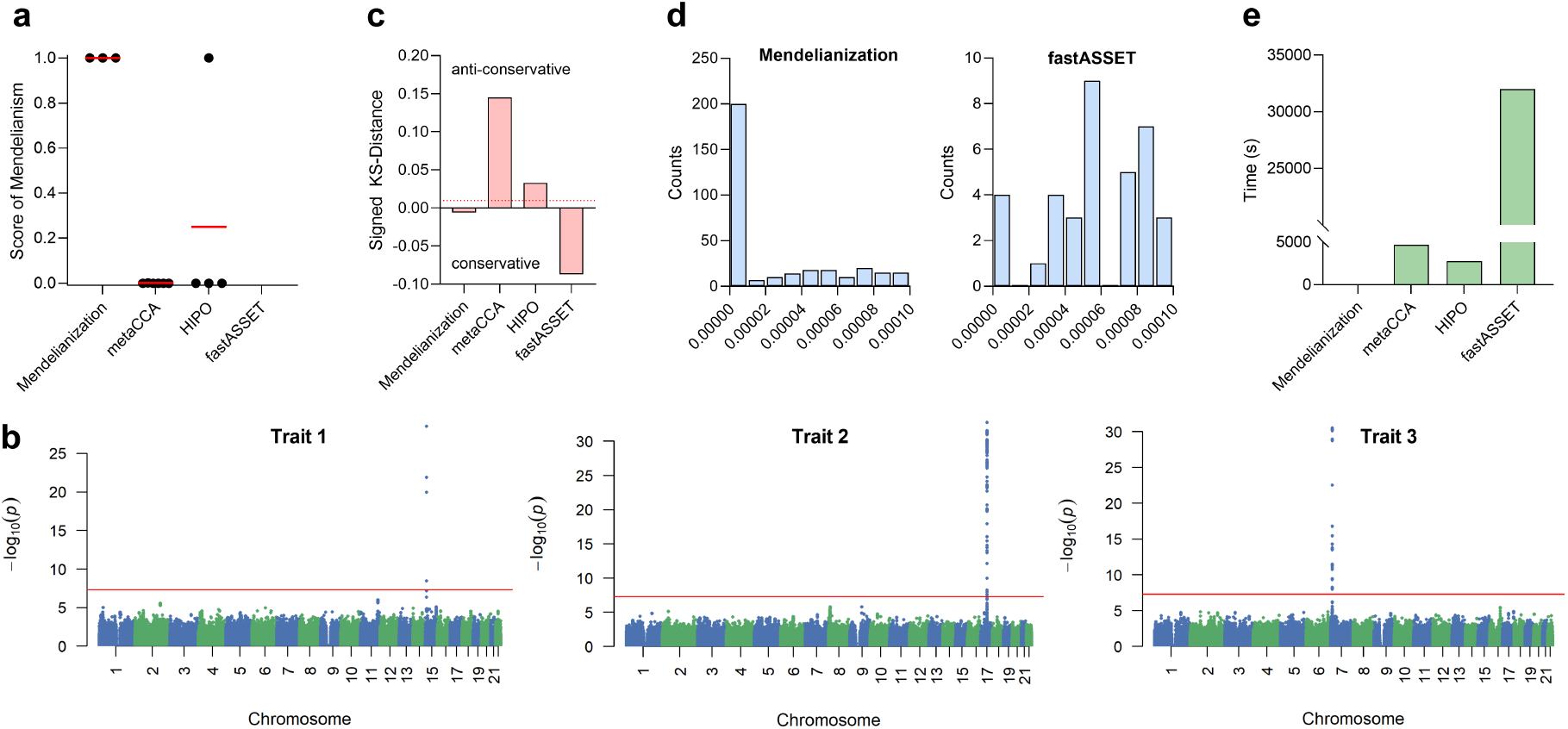
Main results for depression and generalized anxiety. (a) Each dot denotes the SoM for the lead variant within a Manhattan-plot tower, and the red lines correspond to the mean SoMs. Only Mendelianization achieved perfect SoMs for all lead variants. No variants reached the genome-wide significance threshold under fastASSET. (b) For each of the three dots in the Mendelianization column of panel (a), the learned outcome corresponded to an essentially perfect Mendelian-like trait because it produced a Manhattan plot with a single tower, consistent with Theorem 1. (c) Only Mendelianization produced well-calibrated *p*-values; fastASSET was conservative. (d) Among methods with well-calibrated or conservative *p*-values, Mendelianization achieved the highest proxy power by a wide margin. (e) Mendelianization was approximately four orders of magnitude faster than alternative methods.

The chromosome 15 signal with lead variant rs8038726 lies in the 15q11-q13 GABA_A_ receptor cluster (GABRB3, GABRA5, GABRG3), which encodes subunits of the major inhibitory GABA_A_ receptor system. Prior work has implicated GABRB3 and GABRA5 in panic disorder, and broader GABAergic deficits have been linked to major depression and stress-related affective pathology.^10, 14^ The same receptor system also mediates the pharmacological effects of benzodiazepines and related hypnotics.^28^ This annotation is consistent with the first effect-allele-oriented profile, which was characterized by role-impairing irritable anxious distress.

The dense chromosome 17 block with lead variant rs2696455 falls within the structurally complex 17q21.31 inversion region, which contains several neuropsychiatrically relevant genes, including CRHR1, KANSL1, and MAPT. This region should therefore be interpreted as a multi-gene regional signal. Consistent with this view, transcriptome-wide and genetic analyses of anxiety- and stress-related psychiatric phenotypes have implicated multiple genes and transcripts in this region.^31, 17^ CRHR1 nevertheless provides a plausible stress-system annotation because it encodes a corticotropin-releasing hormone receptor involved in HPA-axis regulation, and CRHR1 variation has been linked to depression-relevant phenotypes, including gene-environment interactions involving childhood maltreatment and variation in HPA-axis reactivity.^35^ In our analysis, this locus corresponded to a specific fatigue-weight-gain persistent-worry phenotype.

The chromosome 7 locus with lead variant rs1990622 overlaps TMEM106B, which multiple GWAS and transcriptomic analyses have implicated in depression risk, depression with anxiety, and related symptomatology.^12, 6^ Experimental work has also linked TMEM106B perturbation to depression-like and anxiety-like phenotypes in mice.^12, 18^ This locus corresponded to the third profile, episodic weight-loss depression. Overall, the three loci thus localize to biologically plausible candidate regions whose annotations are consistent with the locus-specific coefficient profiles recovered by Mendelianization.

We next evaluated secondary performance metrics. Only Mendelianization produced well-calibrated *p*-values (Figure 3 (c)). FastASSET generated conservative *p*-values, whereas metaCCA and HIPO yielded anti-conservative estimates, indicating unreliable significance levels. We plot the individual histograms in Supplementary Figure 6. Among methods with calibrated or conservative behavior, Mendelianization achieved the greatest proxy power by far (Figure 3 (d)). The algorithm also required only 2 seconds to complete, while the other methods were slower by at least four orders of magnitude (Figure 3 (e)). Overall, Mendelianization uniquely combined well-calibrated inference, high proxy power, and rapid computation. Finally, ablation results confirmed the necessity of all components of the algorithm (Supplementary Figure 7).

### Alcohol Use Disorder

We increased the difficulty in alcohol use disorder by running the algorithms with only 10 outcome variables corresponding to all individual items of the Alcohol Use Disorders Identification Test (AUDIT).^25^ We again used European ancestry summary statistics from the Pan-UK Biobank with the same quality control. According to Theorem 1 and the simulation results, Mendelianization is unlikely to recover near-perfect Mendelian-like traits with only 10 outcomes, but we still expect it to find approximate Mendelian-like traits. The condition number of Γ was 10.5, indicating that the matrix was well-conditioned for inversion.

The results, summarized in Figure 4, support this expectation. Mendelianization did not achieve perfect SoMs, but every locus achieved higher scores than those from any competing method. Visualization of the per-outcome genome-wide Manhattan plots revealed a substantial, though not perfect, degree of Mendelianism (Figure 4 (b)). Each trait was associated with an interpretable set of canonical coefficients (Supplementary Figure 8).

**Fig. 4.**
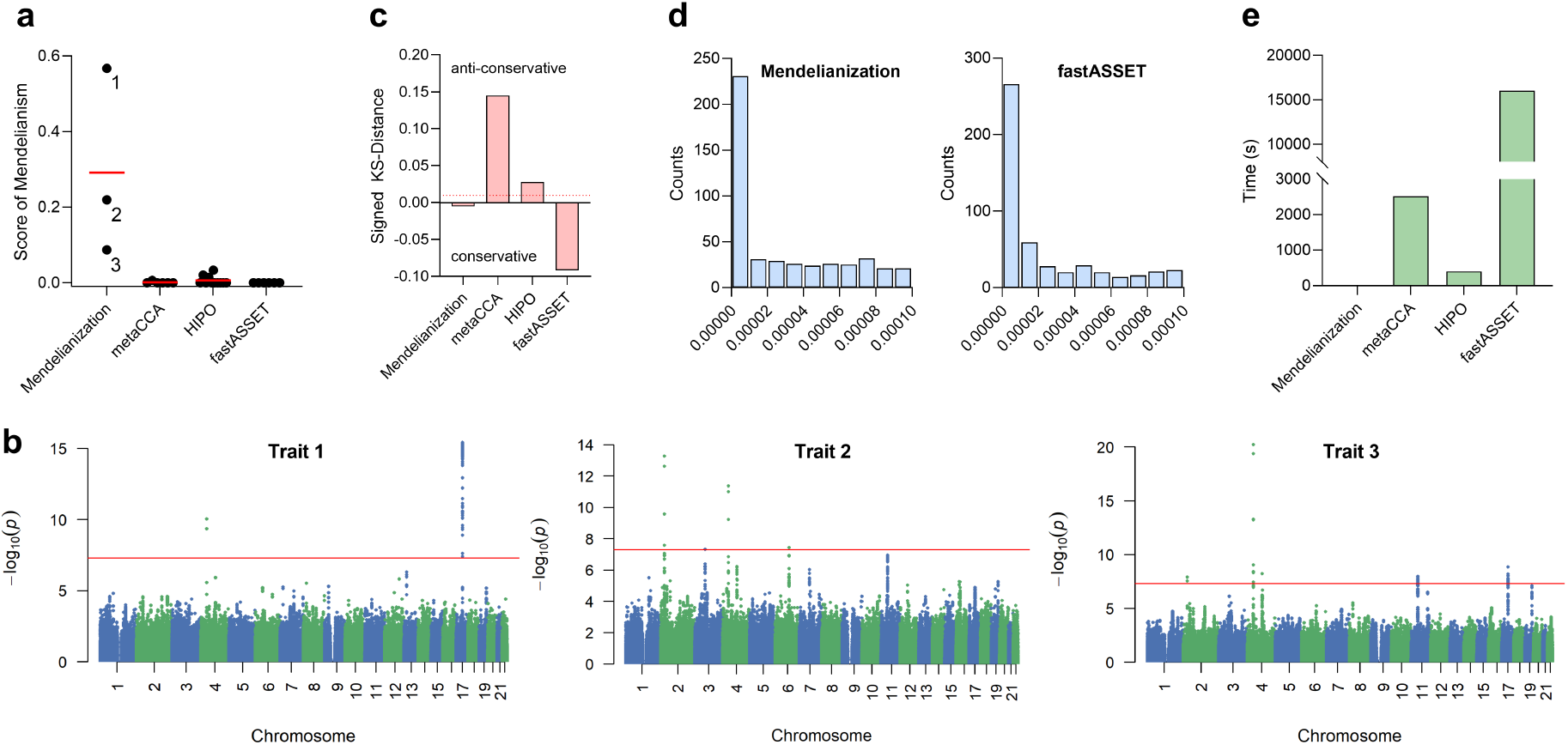
Main results for alcohol use disorder. (a) Mendelianization achieved the highest SoMs for each lead variant. (b) The learned outcome for each point in the Mendelianization column of panel (a) approximated a Mendelian-like trait. (c) Only Mendelianization produced well-calibrated *p*-values. (d) Both Mendelianization and fastASSET attained high proxy power. (e) Mendelianization completed in 0.58 seconds.

The three loci discovered by Mendelianization also localize to biologically plausible alcohol-related regions. The chromosome 17 tower with lead variant rs41382552 again lies in the structurally complex 17q21.31 inversion region encompassing CRHR1–MAPT. This signal should be interpreted as regional rather than as definitively CRHR1-specific, but CRHR1 again provides a plausible stress-axis annotation: CRHR1 variation has been associated with high alcohol intake, binge drinking, and alcohol-dependence-related phenotypes, and has also been studied in interaction with traumatic or stressful life exposure.^34, 23^ The annotation is thus consistent with the first effect-allele-oriented AUD profile, which was characterized by higher alcohol-related consequence components despite lower drinking-frequency and heavy-consumption components, suggesting a stress- or negative-affect-related vulnerability to alcohol-related consequences rather than a simple high-exposure drinking signal. The chromosome 2 signal with lead variant rs780093 maps to the GCKR region, where correlated variants have been robustly associated with alcohol consumption and AUDIT scores in large GWAS.^24^ Functional work further supports a role for this metabolic locus in alcohol-related phenotypes: because C. elegans lacks a direct GCKR ortholog, Thompson et al. perturbed glucokinase as the downstream effector and found altered ethanol-response phenotypes,^33^ while GCKR P446L knock-in mouse work supports functional consequences of the human GCKR alcohol-associated variant.^15^ This locus corresponded to the second profile, characterized by socially noticed regular drinking without the strongest injury or loss-of-control components. The chromosome 4 locus with lead variant rs11940694 overlaps KLB, encoding *β*-Klotho, the FGF21 co-receptor. Human GWAS and experimental studies converge on the FGF21–KLB pathway as a regulator of alcohol preference and intake: brain-specific *β*-Klotho disruption increases alcohol preference in mice, whereas FGF21 signaling suppresses alcohol drinking in rodents and non-human primates.^27, 37, 7^ This locus corresponded to the third profile, characterized by high-frequency drinking with preserved role function, consistent with a mechanism acting primarily on alcohol preference or intake rather than on severe functional impairment. Overall, external genetic and experimental evidence supports locus-level roles for 17q21.31/CRHR1, GCKR, and KLB in distinct alcohol-related dimensions.

As before, Mendelianization was the only method to yield well-calibrated *p*-values (Figure 4 (c); Supplementary Figure 9), while both Mendelianization and fastASSET achieved high proxy power (Figure 4 (d)). Finally, Mendelianization completed at least three orders of magnitude faster than competing methods (Figure 4 (e)), and ablation results confirmed the necessity of the individual components of the algorithm (Supplementary Figure 10). We conclude that Mendelianization was again the only method that simultaneously achieved high degrees of Mendelianism, well-calibrated inference, high proxy power, and rapid computation.

## Discussion

Mendelianization learns composite outcomes whose associations asymptotically concentrate at a single locus. The locus contains at least one causal variant under just four structural assumptions – low-dimensional confounding, outcome diversity, signal floors, and isolation-spillover. The algorithm also handles partial sample-overlap, yields cross-outcome comparable canonical coefficients, and quantifies single-locus concentration via the SoM. In short, Mendelianization performs genome-wide, locus-level causal inference using a substantially modified one-dimensional CCA.

In practice, the outcome set should be chosen to match the scientific question. We recommend moderate, targeted collections of outcomes that densely measure a heterogeneous domain, such as symptom items within a disorder, because the resulting composites remain interpretable as locus-specific profiles within that domain. Phenome-wide outcome sets are possible in principle, but they are not the default use case we recommend. They can be harder to interpret, and at fixed sample size, increasing *m* also increases the degrees of freedom of the 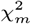 test, so power need not improve unless the added outcomes carry relevant signal. Large outcome sets may also create two separate practical issues: finite-sample calibration of the 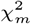 test can deteriorate if Γ is poorly estimated, and the learned latent outcomes may become less stable.

Mendelianization is best suited to harmonized datasets in which outcome definitions, ancestry composition, covariate adjustment, and sample-overlap structure are reasonably consistent across outcomes. In heterogeneous meta-analytic summary statistics, differences in study design, ancestry, imputation, covariate adjustment, sample overlap, or phenotype definition may distort the empirical cross-outcome null covariance of the genome-wide *z*-statistics, summarized by Γ, and affect calibration or interpretation. We therefore recommend harmonizing phenotypes and covariates before analysis, or applying Mendelianization separately within each cohort and evaluating whether the same prespecified procedure recovers similar locus-anchored constructs and localization patterns. Following the recommendations of task-aligned outcome learning,^29^ fixed learned weights should be transferred directly only to closely matched datasets.

In downstream biological or clinical studies, the learned coefficient profile provides a hypothesis about the symptom construct most specifically associated with effect-allele dosage of the lead variant at a given locus. Investigators could thus use the weighted composite directly as a refined locus-specific outcome, or use the coefficient pattern to design a cleaner phenotype that more directly measures the inferred construct. For example, if a learned profile indicates that higher effect-allele dosage at a lead variant is associated with role-impairing irritable anxiety, follow-up studies could use the weighted symptom score or measure that construct more directly. Once this locus-anchored phenotype is defined, standard downstream analyses – fine-mapping, functional annotation, pathway analysis, cellular or animal-model experiments, and pharmacologic follow-up – can proceed as they would for any fixed GWAS outcome.

Despite these strengths, Mendelianization has limitations. First, it cannot resolve LD within a locus because it is a one-dimensional method; isolating variant-level causality generally requires fine-mapping – a separate class of techniques fundamentally distinct from Mendelianization. Second, Mendelianization cannot accommodate completely non-overlapping samples between outcome pairs, which poses challenges when distinct populations are assessed with different measures. Third, Mendelianization remains a linear method that may miss nonlinear confounding and causal effects. Fourth, the present implementation uses an unregularized inverse of Γ. Although the condition numbers observed in our experiments and real-data applications did not indicate numerical instability, regularization may be useful in settings with many highly redundant or nearly collinear outcomes. Future research should therefore explore combining Mendelianization with fine-mapping approaches, extending it to handle fully non-overlapping samples, generalizing it to nonlinear settings, and developing regularized versions for highly collinear outcome panels.

In summary, Mendelianization provides a powerful framework for isolating causal loci to the extent permitted by available outcomes, advancing a reductionist approach that helps clarify – rather than complicate – the pathophysiological understanding of complex disease.

## Data Availability

All data is publicly available in the Pan-UK Biobank (https://pan.ukbb.broadinstitute.org/).

## Competing interests

No competing interest is declared.

## Author contributions statement

E.V.S. conceived, designed, and executed the project in its entirety.

## Acknowledgments

TBD

## Supplementary Materials

### A. Derivation of CCA Solution

Correlation and, likewise, the solution to Expression (1) is scale invariant, so we can impose the constraint *α*^⊤^Σ*α* = 1 and maximize the numerator *r*_*j*_ *α*. We invoke the Cauchy-Schwarz inequality in the Σ-inner product:

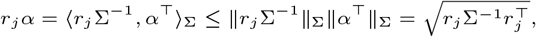

where the last equality follows because we imposed the constraint 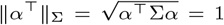. The inequality becomes an equality when 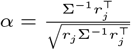. Recall that the solution to Expression (2) is scale-invariant, so we set 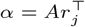 with *A* = Σ.

### B. Finite *m* Assumptions

#### Assumption 5.

*(Low-dimensional latent confounding) For a prespecified m* ∈ ℕ ^+^, *the confounding effects are contained in a subspace U*^*m*^ ⊂ ℝ^*m*^, *and there exists an integer constant r*_0_ ≤ 0 *such that* dim(*U*^*m*^) ≥ *r*_0_. *We additionally assume that each vector of variant-outcome correlations admits a structural decomposition*

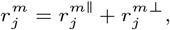

*where* 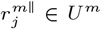 *is the confounding component and* 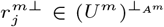 *is the causal component, with orthogonality defined by A*^*m*^ = (Σ^*m*^)^−1^.

#### Assumption 6.

*(Outcome diversity) We have* 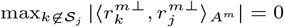 *for a prespecified m*.

#### Assumption 7.

*(Signal floor) For a prespecified m, if l* ∈ *S*_*j*_ *causes* 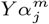, *then there exists a constant β*_c_ *>* 0 *such that* 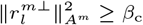. *Moreover, there exists a constant β*_on_ *>* 0 *such that* 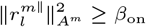 *for every l* ∈ *S*_*j*_ *that does not cause* 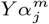.

#### Assumption 8.

*(Isolation-spillover) If some l* ∈ *S*_*j*_ *causes* 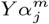, *we have* 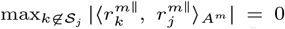 *for a prespecified m. On the other hand, if there does not exist l* ∈ *S*_*j*_ *that causes* 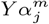, *then* 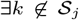 *with* 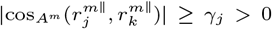 *and* 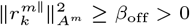 *for a prespecified m*.

### C. Pseudocode & Time Complexity Analysis

We present the pseudocode for Mendelianization in Algorithm 1. The algorithm first computes *Ŝ* in Line 1, which estimates the realized conditional covariance Γ^(*n*)^(*M*) under Theorem 2 and targets Γ in the large-sample limit. Using this estimate, it then derives the raw canonical coefficients in Line 2 by solving Expression (5). These coefficients also form the basis of the *Q* statistics in Expression (3) that enable hypothesis testing; recall that each *Q*_*j*_ follows a 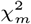 distribution under the null, so the algorithm can readily compute *p*-values in Line 4. Next, Mendelianization computes SoMs for lead variants that surpass a genome-wide significance threshold (e.g., 5 × 10^−8^) in Line 5. Finally, the algorithm converts the raw coefficients into their interpretable form *α*′ by applying Proposition 4 in Line 6. Thus, although the theory of Mendelianization is intricate, the resulting algorithm is remarkably straightforward.

#### Algorithm 1

Mendelianization

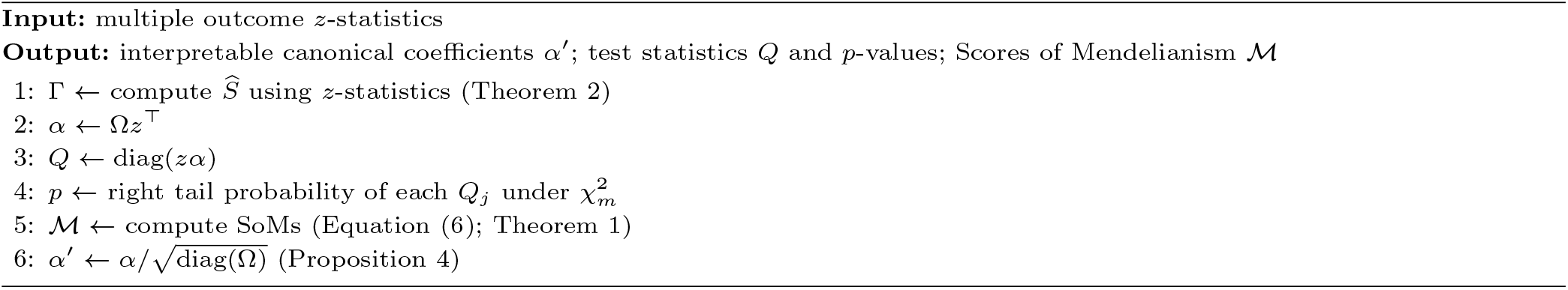

The algorithm also has a favorable time complexity. In particular, estimating the correlation matrix Γ described in Line 1 costs *O*(*qm*^2^) time. In Line 2, inverting Γ requires *O*(*m*^3^) time, and the matrix product Ω*z*^⊤^ required to estimate *α* costs *O*(*m*^2^*q*). Constructing all *Q*_*j*_ statistics and evaluating their *p*-values in Lines 3 and 4 adds *O*(*mq*), and computing *α*′ in Line 6 requires another *O*(*mq*). For the SoM step, suppose there are *s* significant variants, and each constitutes its own tower to take the worst case. Computing each statistic Z_*k*|*j*_ requires *O*(*m*) time for the numerator and *O*(*m*^2^) for the denominator in the definition of Z_*k*|*j*_. Moreover, the numerator must be evaluated genome-wide over *q* variants, so an upper bound on the total cost of SoM amounts to *O*(*s*(*mq* + *m*^2^)). Summing these components, the overall complexity of Mendelianization is *O*(*m*^3^ + *m*^2^*q* + *sqm*), where the simplification uses *q* ≫ *m*. Thus, the algorithm scales linearly with the number of variants and cubically with the number of outcome variables. In practice, Mendelianization completes within seconds and is at least three orders of magnitude faster than the competing methods.

### D. Proofs

#### Theorem 1.

- *There exists a constant c*_*j*_ *>* 0 *such that* 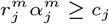 *for sufficiently large m, and* 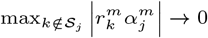, *so Mendelianism is achieved asymptotically*.
- *For all sufficiently large m, there exists at least one variant l*_*m*_ ∈ *S*_*j*_ *that causes* 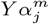.

*Proof* We first prove the forward direction by contrapositive. Suppose it is not the case that, for all sufficiently large *m*, there exists a variant *l* ∈ *S*_*j*_ that causes 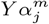. Thus, for every *M*_1_ *>* 0, there exists *m* ≥ *M*_1_ such that no variant *l* ∈ *S*_*j*_ causes 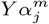. As a result, *j* ∈ *S*_*j*_ does not cause 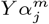. We need to show 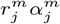 is not bounded below by a positive constant for sufficiently large *m* or 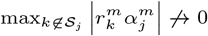 (or both). We will show that 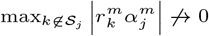.

Assumption 1 allows us to write:

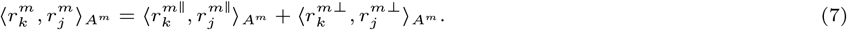

Let 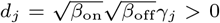 and 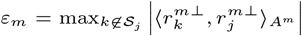. Under Assumption 2, *ε*_*m*_ → 0, so there exists *M*_2_ *>* 0 such that for all *m* ≥ *M*_2_, *ε*_*m*_ *< d*_*j*_ /2 and Assumptions 3 and 4 apply.

Choose any *M*_1_ ≥ *M*_2_. By the supposition above, there exists *m* ≥ *M*_1_ such that no variant *l* ∈ *S*_*j*_ causes 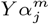. By Assumptions 3 and 4, there exists *k*∉ *S*_*j*_ such that

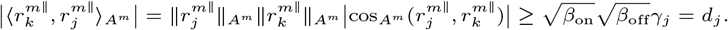

The reverse triangle inequality allows us to write:

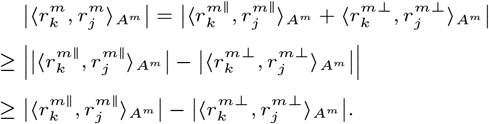

As a result,

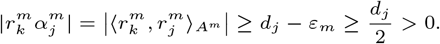

Since *M*_1_ ≥ *M*_2_ was arbitrary, for every sufficiently large *M*_1_ there exists *m* ≥ *M*_1_ such that

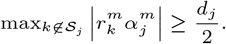

Therefore, lim 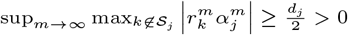, so the off-locus associations cannot converge to zero. This completes the proof of sufficiency – i.e., asymptotic Mendelianism is sufficient to establish causality.

To prove the backward direction, assume that, for all sufficiently large *m*, there exists at least one variant *l*_*m*_ ∈ *S*_*j*_ that causes 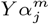. Then, we can invoke Assumption 1 and write Equation (7) for any *k*. We have two situations:

1. If *k*∉ *S*_*j*_, then

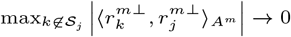

by Assumption 2. Moreover,

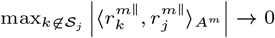

by the first clause of Assumption 4. Hence,

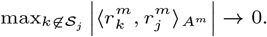
2. If *k* ∈ *S*_*j*_ and *k* = *j*, then we have two situations. If *j* causes 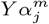, then the first clause of Assumption 3 allows us to write:

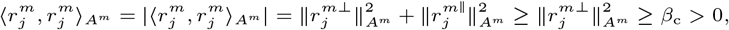

for sufficiently large *m*. On the other hand, if *j* does not cause 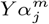, then the second clause of Assumption 3 allows us to write:

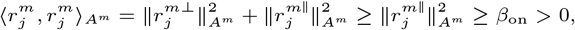

for sufficiently large *m*.

We conclude that there exists a constant *c*_*j*_ = min{*β*_c_, *β*_on_} *>* 0 such that 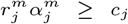 for sufficiently large *m*, and 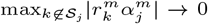, which completes the proof of necessity – i.e., asymptotic Mendelianism is necessary to establish causality.

□

#### Proposition 1.

i. *We have* 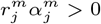 *and* 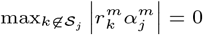, *so Mendelianism is perfectly achieved for S*_*j*_ *at m*.
ii. *At least one variant l* ∈ *S*_*j*_ *causes* 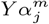.

*Proof* The proof almost exactly parallels that of Theorem 1, but we include it here for completeness. We first prove the forward direction by contrapositive. Suppose there does not exist a variant *l* ∈ *S*_*j*_ that causes 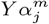. As a result, *j* ∈ *S*_*j*_ does not cause 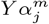. We need to show 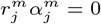 or 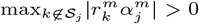 (or both) for prespecified *m*. We will show that 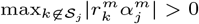.

Assumption 5 allows us to write:

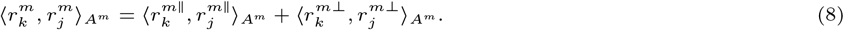

By Assumptions 7 and 8, there exists *k*∉ *S*_*j*_ where:

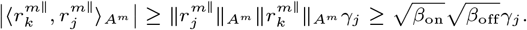

The reverse triangle inequality allows us to write:

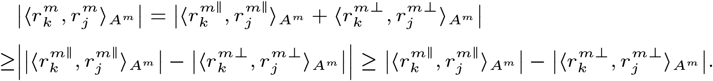

Thus, under Assumption 6, we have:

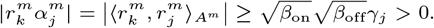

This completes the proof of sufficiency – i.e., perfect Mendelianism is sufficient to establish causality.

To prove the backward direction, assume that there exists at least one variant *l* ∈ *S*_*j*_ that causes 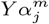. Then, we can invoke Assumption 5 and write Equation (8) for any *k*. We have two situations:

1. If *k*∉ *S*_*j*_, then 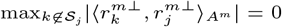 by Assumption 6. Moreover, 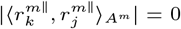 by the first clause of Assumption 8. Hence, 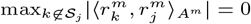.
2. If *k* ∈ *S*_*j*_ and *k* = *j*, then we have two situations. If *j* causes 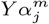, then the first clause of Assumption 7 allows us to write:

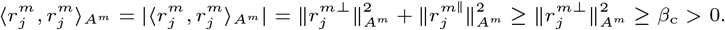

On the other hand, if *j* does not cause 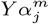, then the second clause of Assumption 7 allows us to write:

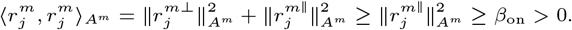

We conclude that 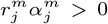 and 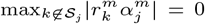, which completes the proof of necessity – i.e., perfect Mendelianism is necessary to establish causality. □

#### Proposition 2.

*Assume i*.*i*.*d. samples across individuals, V* ⫫ *Y, and V* ⫫ *M* | *Y*. *Then* 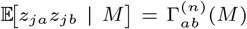, *where* 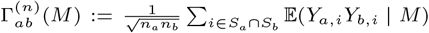. *Separately, without requiring V* ⫫ *Y or V* ⫫ *M* | *Y, under i*.*i*.*d. missingness, with p* = *P* (*M* = 1) *>* 0, *p* = *P* (*M* = 1) *>* 0, *and p* = *P* (*M* = 1, *M* = 1) *>* 0, 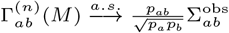.

*Proof* Note that *z*-statistics are computed on observed samples, so

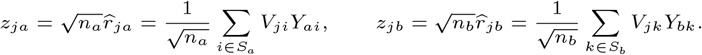

Therefore

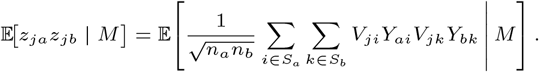

The cross-terms with *i*≠ *k* vanish by i.i.d. sampling and the null assumptions. Indeed, *V* ⫫ *Y* and *V* ⫫ *M* | *Y* imply *V* ⫫ (*Y, M*) by contraction, and hence *V* ⫫ *Y* | *M* and *V* ⫫ *M*. Since the genotypes are standardized, this gives E(*V*_*ji*_ | *M*) = 0. Thus only the terms with *i* = *k* ∈ *S*_*a*_ ∩ *S*_*b*_ remain:

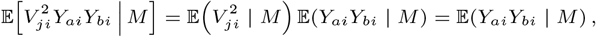

Using *V* ⫫ *Y* | *M* and *V* ⫫ *M*,

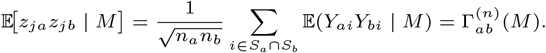

because 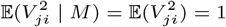. Hence

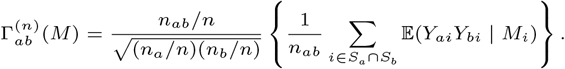

It remains to identify the large-sample limit. Write *M*_*i*_ for the missingness pattern of individual *i*. Under i.i.d. missingness, E(*Y*_*ai*_*Y*_*bi*_ | *M*) = E(*Y*_*ai*_*Y*_*bi*_ | *M*_*i*_), and

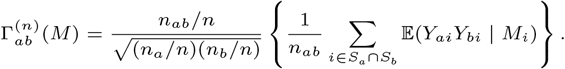

By the strong law of large numbers, *n*_*a*_*/n* → *p*_*a*_, *n*_*b*_*/n* → *p*_*b*_, and *n*_*ab*_*/n* → *p*_*ab*_ almost surely. Also,

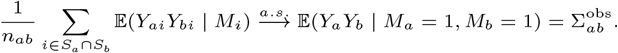

Therefore 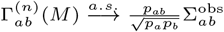. □

#### Proposition 3.

*Assume i*.*i*.*d. samples across individuals, p*_*a*_ *>* 0, *p*_*b*_ *>* 0, *p*_*ab*_ *>* 0, *and the corresponding conditional product moments are uniformly integrable. Then* 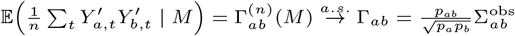 and 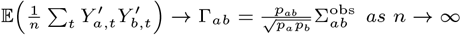.

*Proof* For the first statement, using 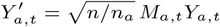, we have

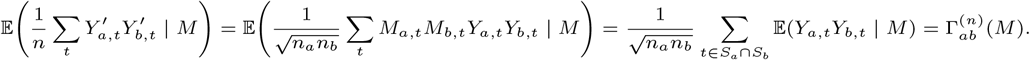

Under i.i.d. missingness,

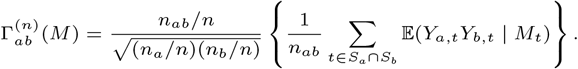

Since *p*_*a*_ *>* 0, *p*_*b*_ *>* 0, and *p*_*ab*_ *>* 0, the strong law of large numbers gives *n*_*a*_*/n* → *p*_*a*_, *n*_*b*_*/n* → *p*_*b*_, and *n*_*ab*_*/n* → *p*_*ab*_ almost surely, and the pairwise-observed average satisfies

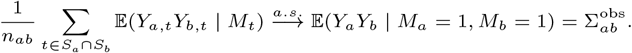

Therefore 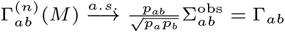.

For the second statement, by the law of total expectation,

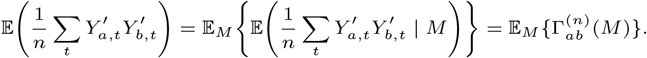

Since 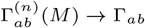 almost surely, and the corresponding conditional product moments are uniformly integrable, it follows that, 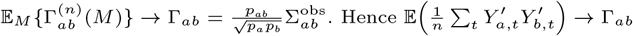. □

#### Theorem 2.

*For fixed outcomes a and b, consider the estimator* 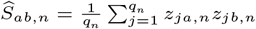, *where q*_*n*_ → ∞ *as n* → ∞. *Let G*_*n*_ *index the variants that are independent of Y, and let B*_*n*_ *index the variants that are dependent on Y*. *Define X*_*j,n*_ = *z*_*ja,n*_*z*_*jb,n*_. *Assume the conditions of the second part of Proposition 2 hold and further assume:*

1. *(Asymptotic null centering) There exists a sequence η*_*n*_ → 0 *such that* 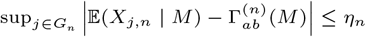
2. *(Vanishing aggregate contamination) We have* 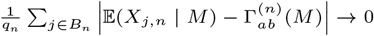
3. *(Local dependence) Conditional on M, the variables* 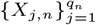 *admit an undirected dependency graph with maximum degree D*_*n*_, *and* 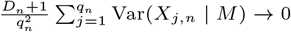.

*Then* 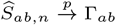.

*Proof* Let *X*_*j,n*_ = *z*_*ja,n*_*z*_*jb,n*_. Consider the unnormalized sum 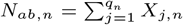, so that 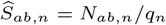. Write:

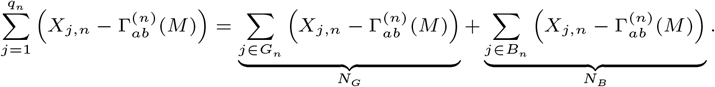

By Condition 1, we have

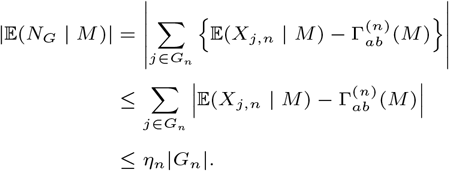

We can also write:

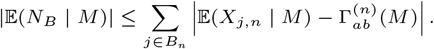

The inequality follows by the triangle inequality. Thus, by Conditions 1 and 2,

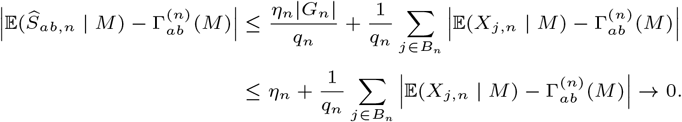

Let *v*_*j,n*_ = Var(*X*_*j,n*_ | *M*), and let *E*_*n*_ denote the edge set of the dependency graph in Condition 3. We can write:

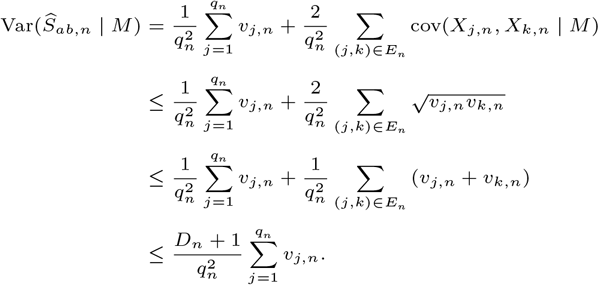

The first inequality follows from the conditional Cauchy–Schwarz inequality, the second from 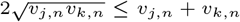, and the final inequality follows because each vertex has degree at most *D*_*n*_. Therefore,Condition 3 implies Var(Ŝ_*ab,n*_ | *M*) → 0.

Let *µ*_*n*_ = E(*Ŝ*_*ab,n*_ | *M*). From the expectation calculation above, 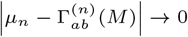. For any *ε >* 0, we therefore have

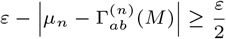

for sufficiently large *n*. We invoke the triangle inequality to write:

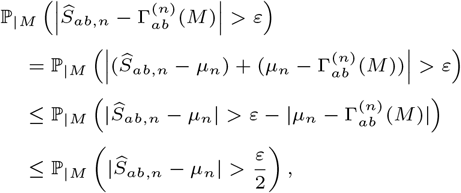

where we have moved the conditioning notation to the subscripts to avoid confusion with absolute value notation. Chebyshev’s inequality gives

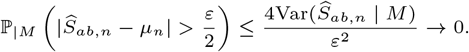

Therefore, 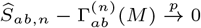 conditional on *M*, and hence also unconditionally by dominated convergence.

Finally, Proposition 2 gives 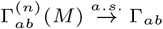 as *n* → ∞. Thus,

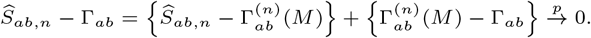

This proves 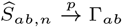. □

### E. Additional Experimental Results

#### Simulations

**Supplementary Fig. 1.**
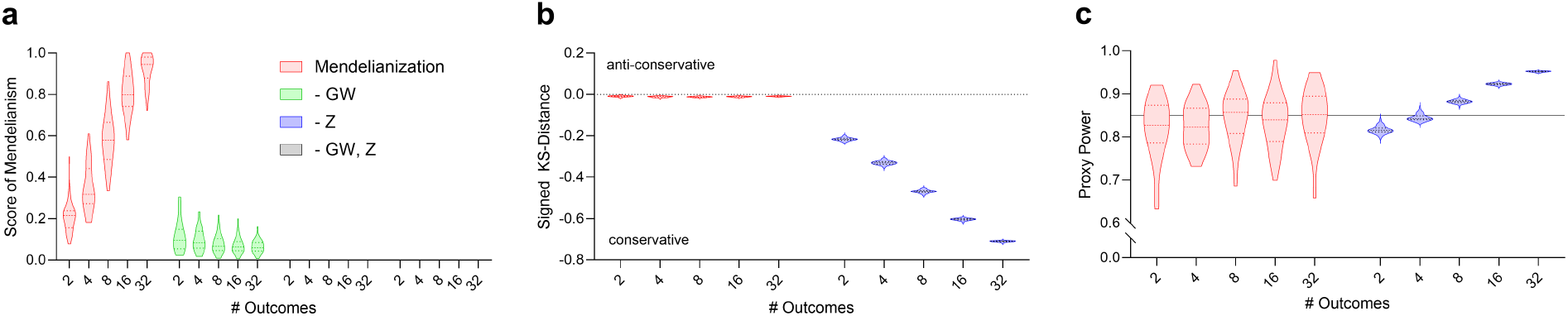
Ablation results. “-GW” denotes not applying each learned outcome genome-wide, “-Z” denotes estimating Γ from *β* coefficients instead of *z*-statistics, and “-GW, Z” denotes using both modifications. (a) Misapplying the CCA-derived outcomes (-GW) or using *β* coefficients in place of *z*-statistics (-Z) reduced the SoM to essentially zero; for two of the ablations, the SoM violin plots collapse at the bottom and are therefore not visible. (b) Using *β* coefficients produced highly conservative *p*-values. (c) The corresponding *p*-value histogram was strongly U-shaped, indicating severe miscalibration and yielding highly unstable power across outcome dimensions. Despite these deficiencies, the -GW and -Z variants still achieved near-perfect true positive rates and completed in under one second, comparable to Mendelianization.

**Supplementary Fig. 2.**
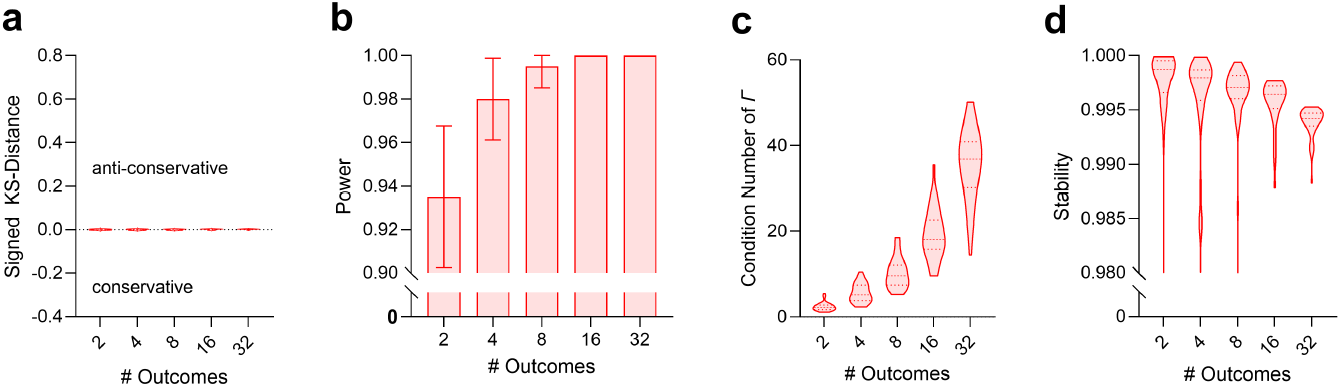
Null-simulation, power, conditioning, and stability results. (a) In global-null simulations, where no variant causally affected any outcome, Mendelianization produced *p*-values that closely followed the expected uniform distribution. (b) In simulations with known causal variants, Mendelianization achieved high empirical power at the genome-wide threshold, with power increasing as the number of outcomes grew. (c) The condition number of Γ increased with the number of outcomes, but never exceeded 51, indicating numerical stability across the simulated settings. (d) Composite stability remained high across all outcome dimensions, with Γ-weighted cosine similarities usually above 0.99. Thus, the increasing condition number did not translate into instability of the learned composites.

#### Simulations with Confounding

To evaluate robustness under latent confounding, we repeated the same semi-synthetic simulation pipeline after adding a sparse low-rank confounded component to the latent traits. Let 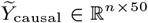 denote the same causal latent-trait matrix generated in the non-confounded simulation from the five causal variants. In each replicate, we additionally sampled 10 LD blocks and selected one tagging variant from each block. These tagging variants were assigned to *r*_0_ = 2 shared latent confounding factors. For each factor *g*, we formed a confounder score by taking a randomly signed sum of the genotypes of the tagging variants assigned to that factor,

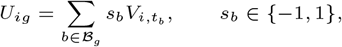

where *t*_*b*_ is the tagging variant selected from confounded block *b*, and then standardized each column of *U*. We then drew a random loading matrix *B*_conf_ ∈ ℝ^2×50^ with independent standard normal entries and generated a raw confounded latent-trait component 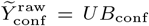. Thus, only two latent confounding factors affected all 50 latent traits, but they did so through heterogeneous trait-specific loadings. To make the confounded component comparable in magnitude to the causal component, we rescaled it as

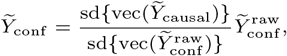

and then generated the final latent traits as 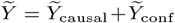. The observed outcomes and summary statistics were then generated from 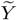 using the same outcome-mixing, sample-size, and blockwise bootstrapping procedure as in the non-confounded simulations.

This construction was designed to mirror the confounding assumptions in the theory. The rank-two matrix *UB*_conf_ corresponds to the low-dimensional latent confounding condition in Assumption 1. The reuse of the same two confounding directions across multiple LD blocks enforces the isolation–spillover behavior in Assumption 4: a purely confounded signal should tend to reappear at other blocks sharing the same latent confounding direction, rather than producing an isolated single-locus tower. Finally, scaling 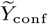 to match the empirical scale of 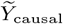 ensures that the confounded component is non-negligible, creating a meaningful stress test rather than a vanishing perturbation.

We summarize the confounded synthetic-data results in Supplementary Figure 3. Overall, Mendelianization produced results that were qualitatively identical to the corresponding no-confounding experiments, supporting the theoretical prediction that low-dimensional spillover confounding should not by itself create isolated Mendelianized towers. The ablation results showed the same pattern (Supplementary Figure 4); Mendelianization was again the only method that simultaneously achieved increasing SoM with more outcomes, maintained Type I error control, and preserved stable proxy power.

**Supplementary Fig. 3.**
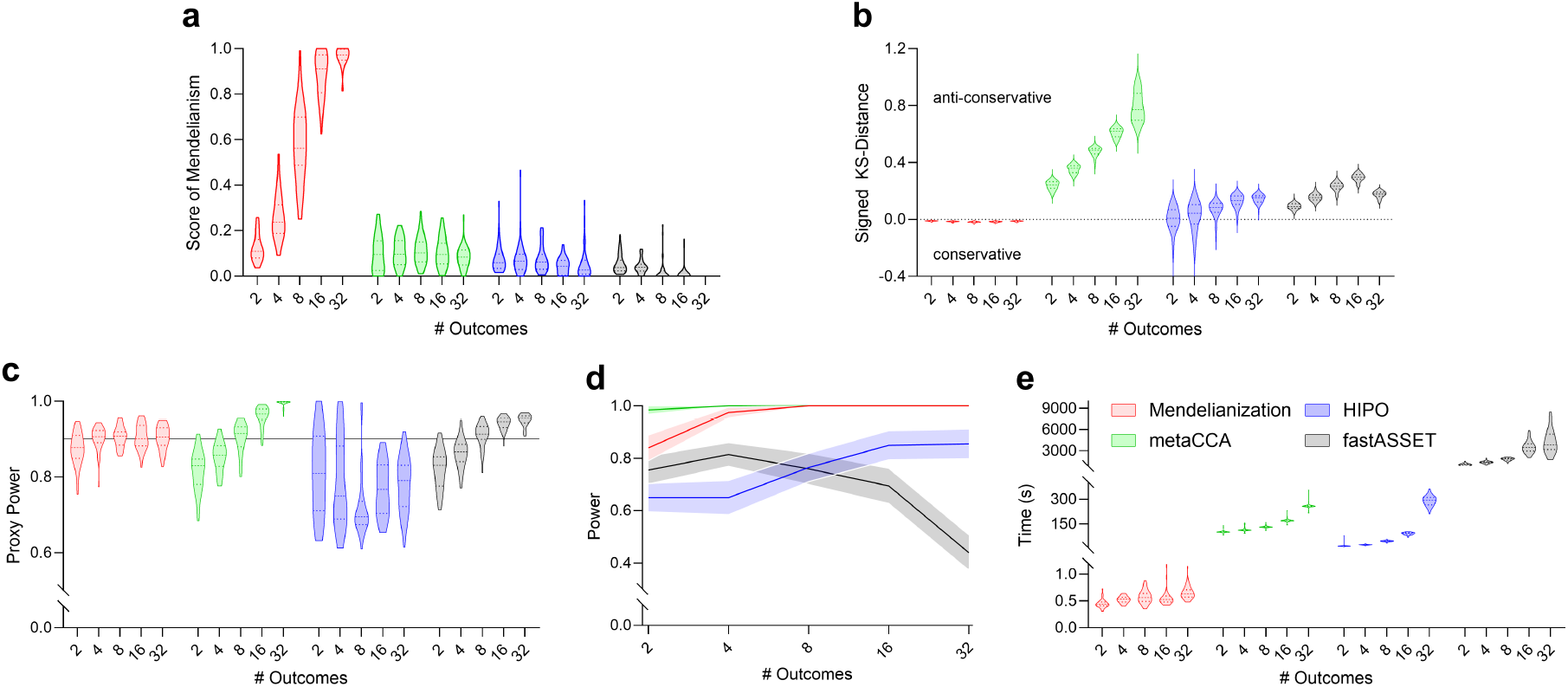
Synthetic-data results with latent confounding. The same simulation design as in Figure 2 was repeated after adding sparse low-rank latent confounding shared across multiple LD blocks. Results closely matched the no-confounding setting.

**Supplementary Fig. 4.**
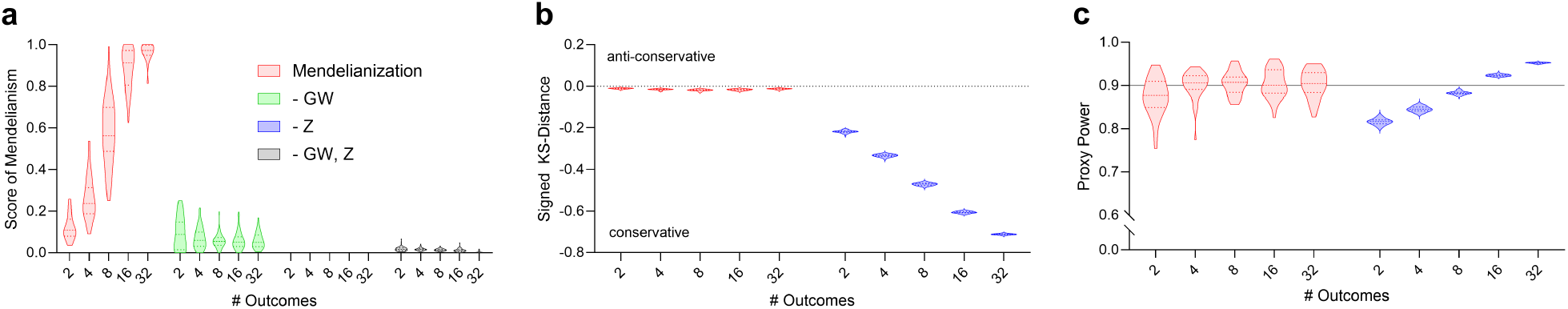
Ablation results with latent confounding. Ablation analyses under the confounded synthetic-data setting qualitatively matched the corresponding no-confounding results in Supplementary Figure 1.

### Depression and Generalized Anxiety

**Supplementary Fig. 5.**
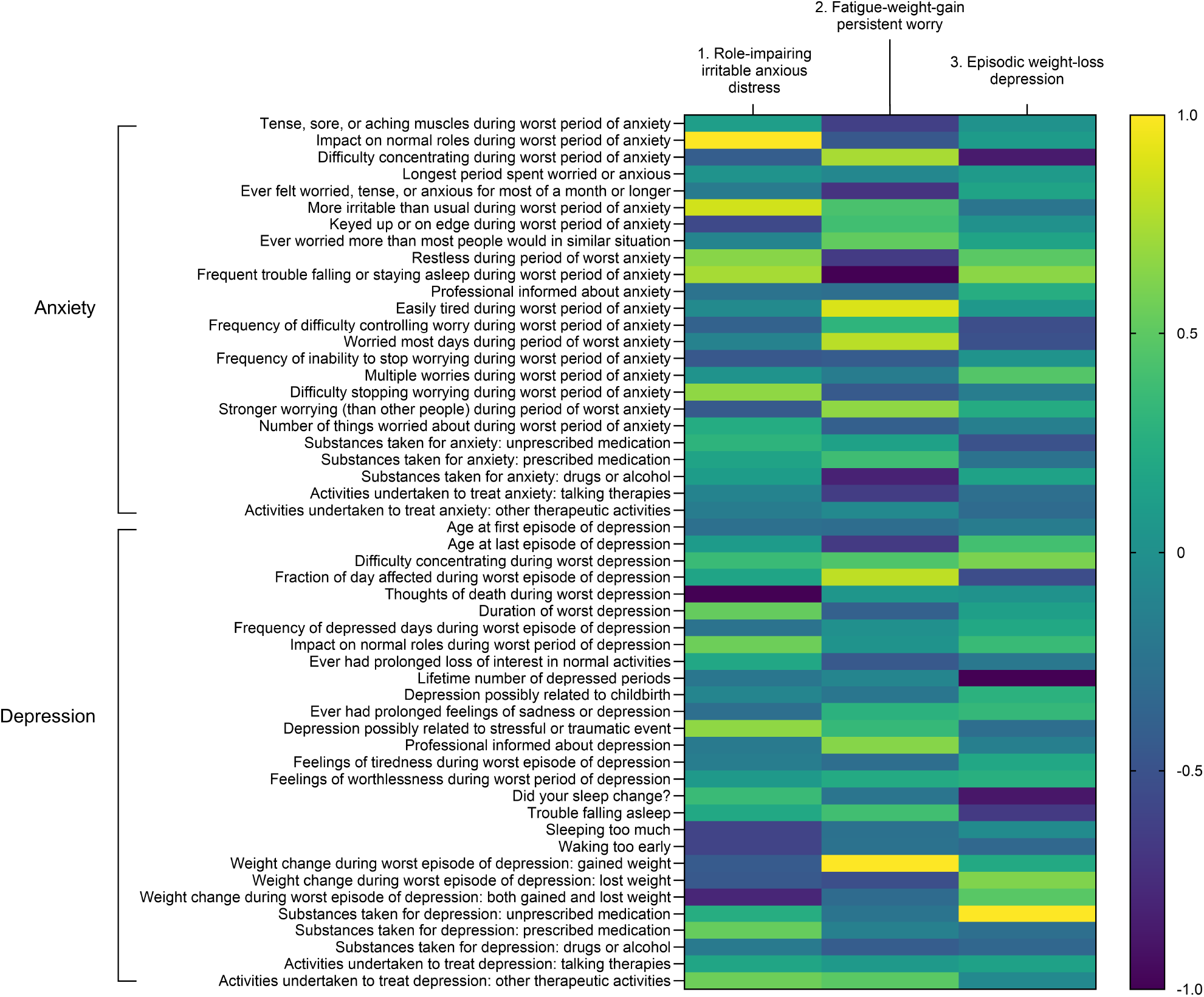
Heatmap of interpretable *α*′ coefficients for lead variants in depression and anxiety. Each column displays the coefficients for a lead variant. The coefficients are interpreted as an allele-oriented symptom profile: positive entries indicate unique symptom components positively associated with higher dosage of the allele used to orient the input *z*-statistics, whereas negative entries indicate unique symptom components negatively associated with higher dosage of that same allele. Thus, the signs should be read jointly within a column as a contrast among residualized symptom components. To emphasize this contrast structure, positive and negative coefficients were normalized separately within each column to the range [*−*1, 1]. The resulting profiles were labeled as: (i) role-impairing irritable anxious distress; (ii) fatigue-weight-gain persistent worry; and (iii) episodic weight-loss depression. These profiles illustrate distinct locus-specific dimensions of depression and anxiety, underscoring the heterogeneity of these disorders.

**Supplementary Fig. 6.**
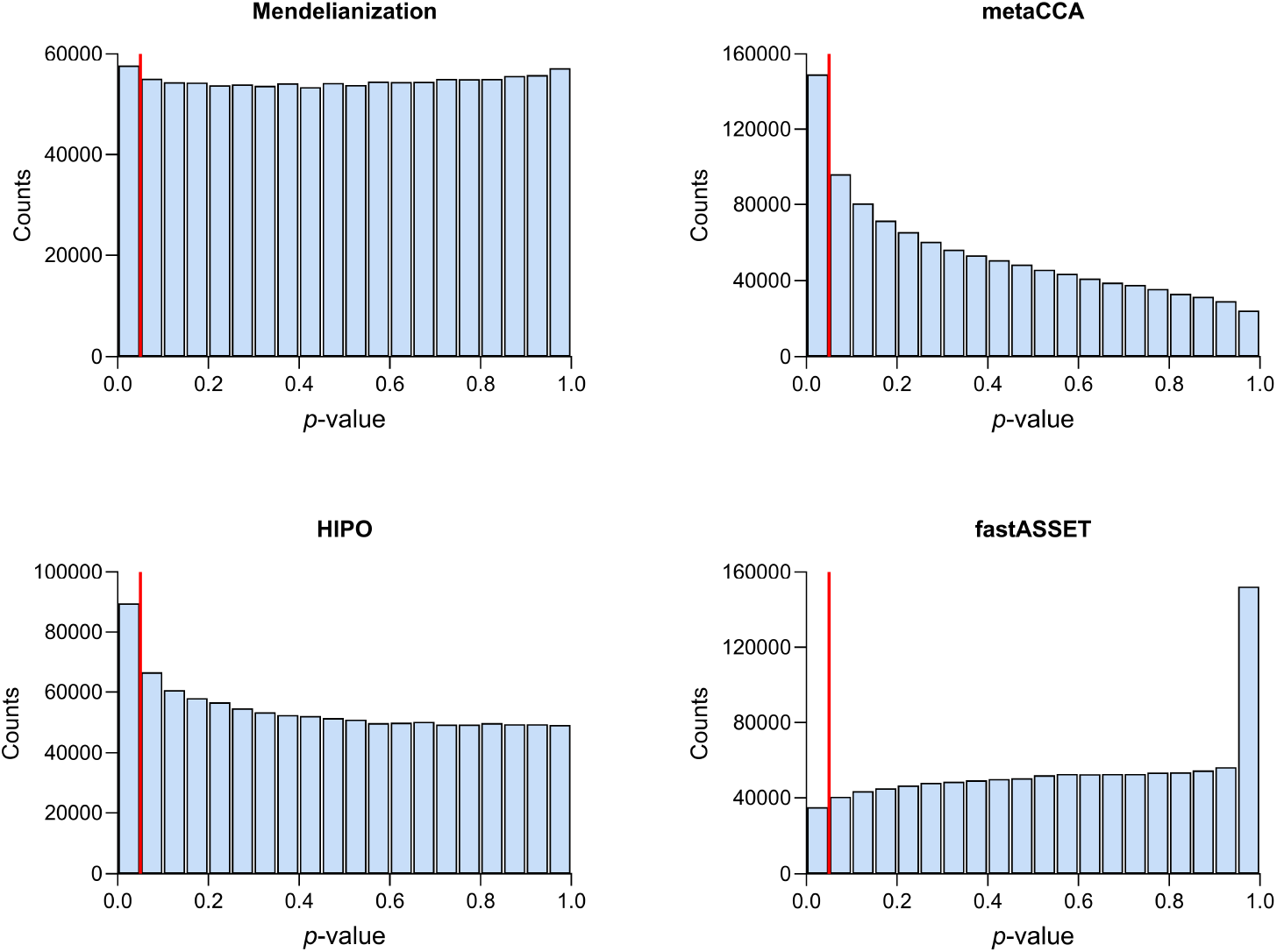
Histograms of *p*-values across all variants. Mendelianization was the only method to produce a near-uniform distribution for variants associated with *p*-values above 0.05 (red line). By contrast, metaCCA and HIPO yielded anti-conservative distributions with shrunken *p*-values, whereas fastASSET was overly conservative, producing *p*-values near one for many variants.

**Supplementary Fig. 7.**
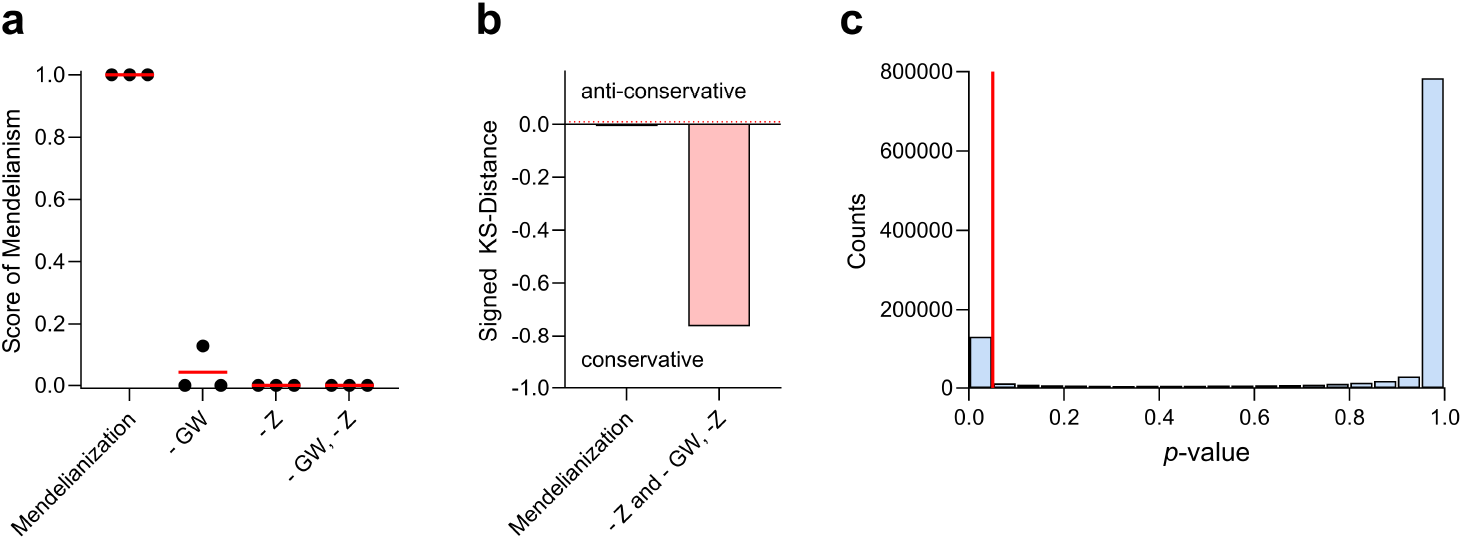
Ablation results. (a) Misapplying the learned outcomes or using *β* coefficients in place of *z*-statistics (or both) decreased the SoM score to near zero. (b) The histogram obtained by *β* coefficients was very conservative. (c) Inspection of the histogram derived from the *β* coefficient estimates revealed highly miscalibrated *p*-values following a U-shape curve.

### Alcohol Use Disorder

**Supplementary Fig. 8.**
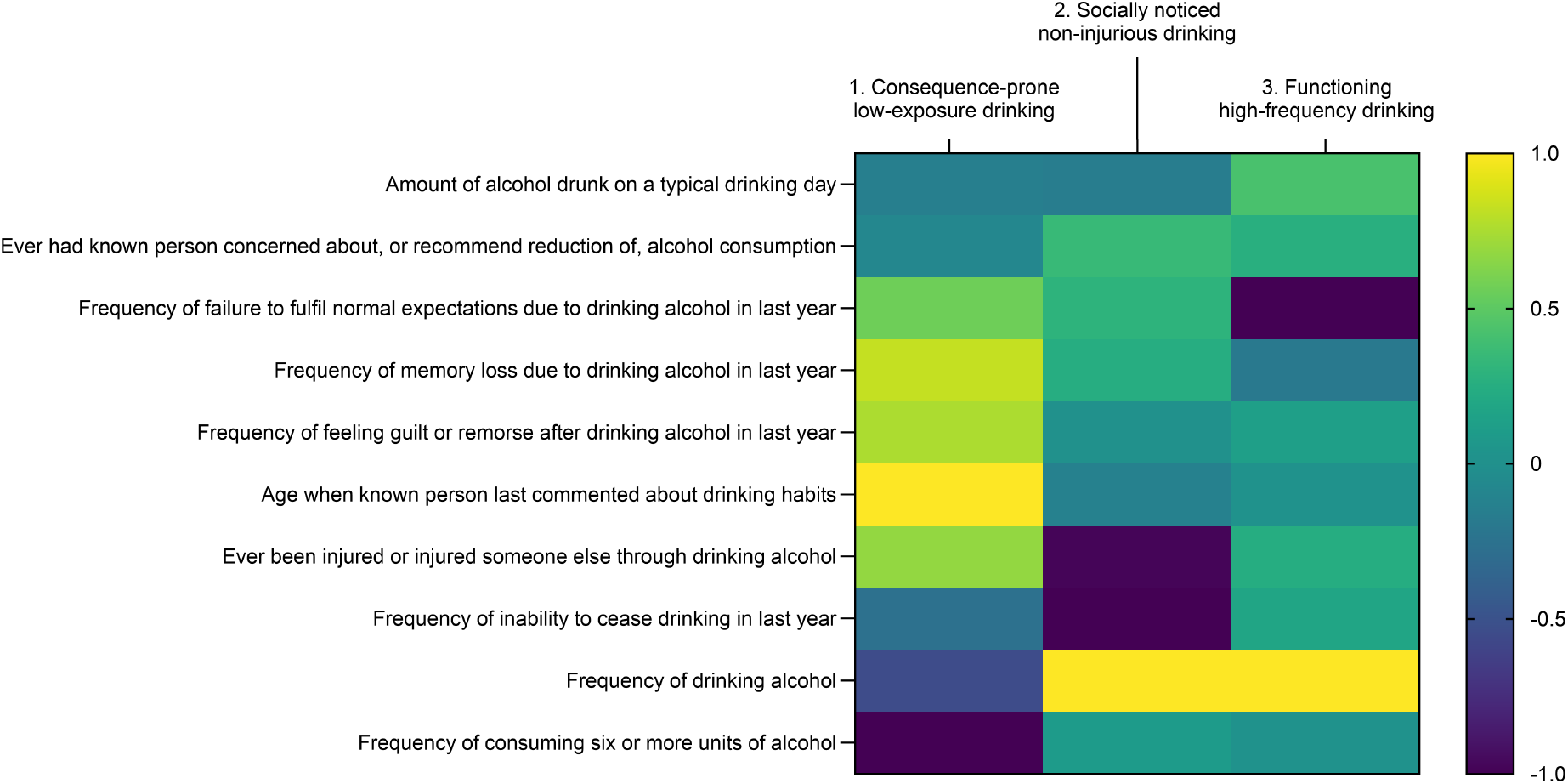
Heatmap of *α*′ coefficients for lead variants in alcohol use disorder. As in Supplementary Figure 5, each column should be read as a signed, allele-oriented profile of unique AUDIT item components. Positive entries identify AUDIT components associated with higher effect-allele dosage at the lead variant, whereas negative entries identify components associated in the opposite direction. The labels summarize the overall profile of each column. The coefficient patterns resolve three AUD profiles: (i) consequence-prone low-exposure drinking; (ii) socially noticed regular drinking without severe complications; and (iii) high-frequency drinking with preserved role function.

**Supplementary Fig. 9.**
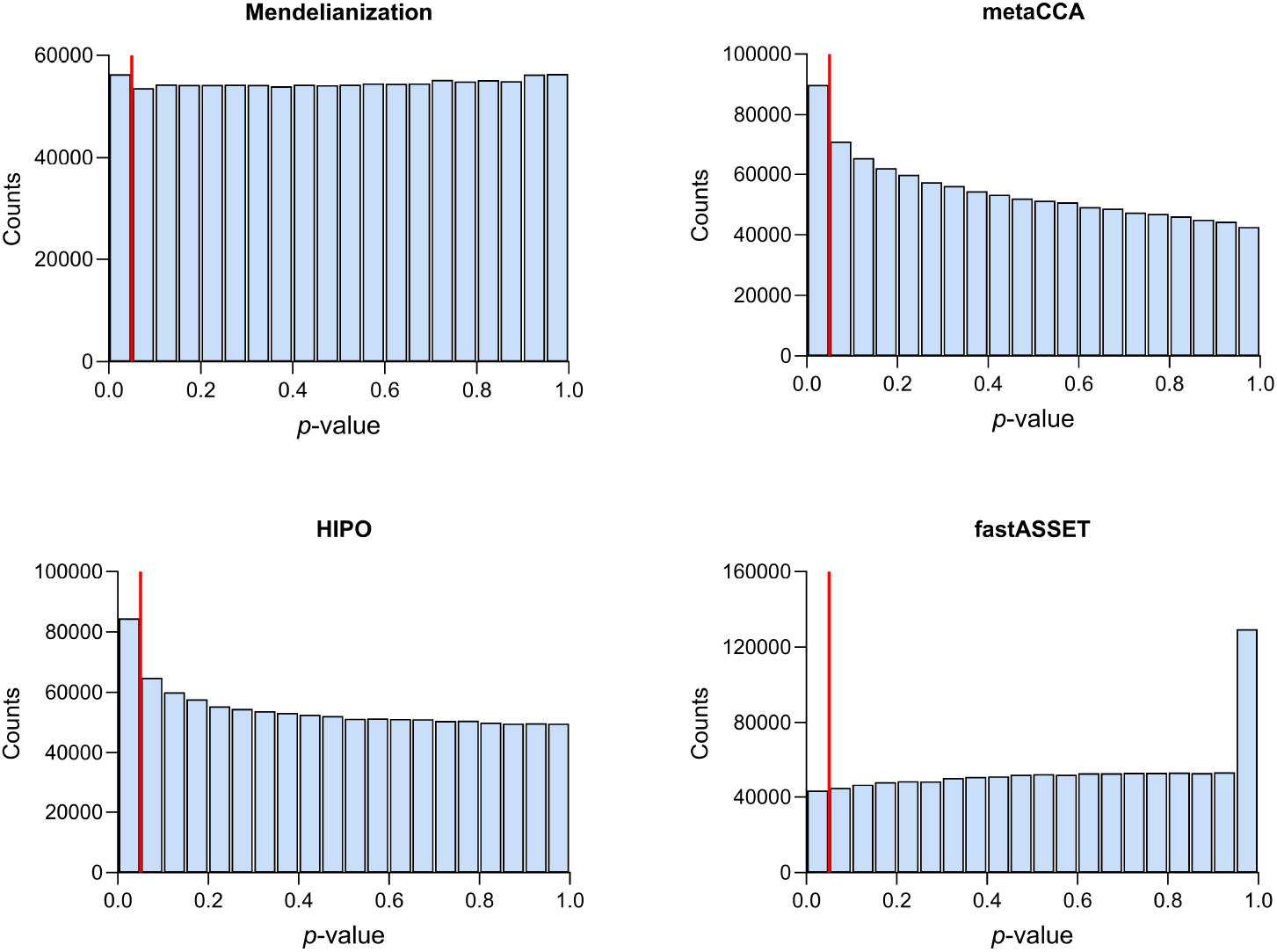
Histograms of *p*-values across all variants. Mendelianization was again the only method to produce a near-uniform distribution on [0.05, 1].

**Supplementary Fig. 10.**
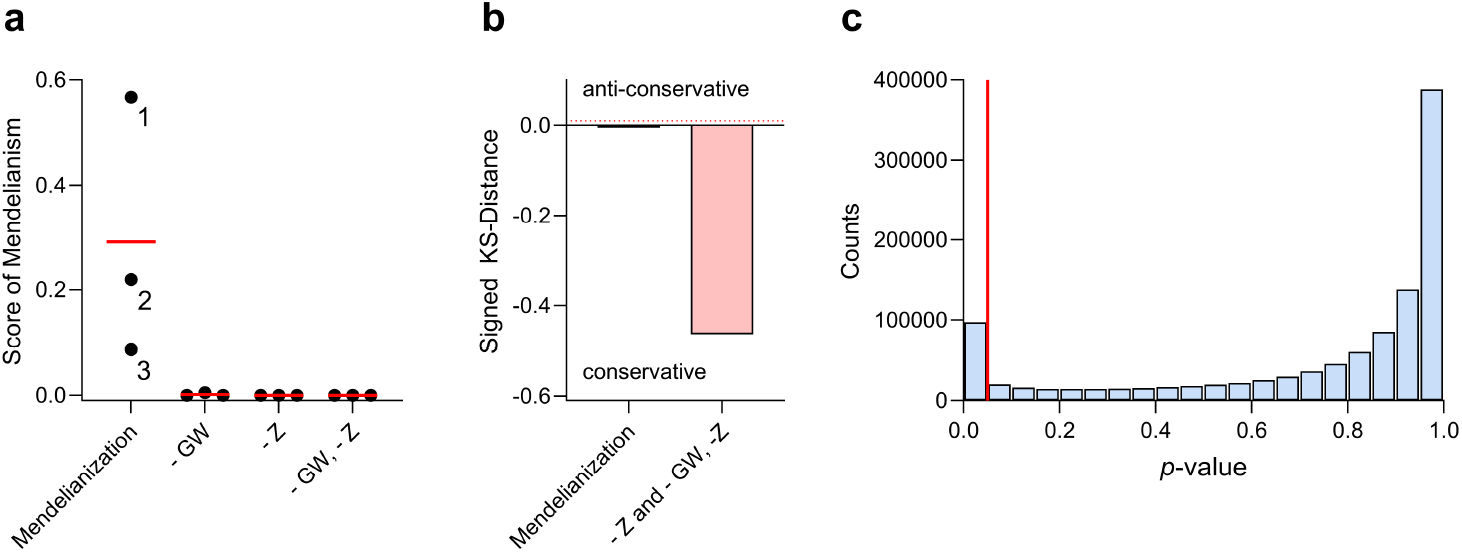
Ablation results. (a) Mendelianization again achieved the highest SoMs, and (b, c) estimating Γ from *β* coefficients yielded highly miscalibrated *p*-values.

## Footnotes

1 All arguments can be modified by substituting sample sizes with Fisher-effective sample sizes in generalized linear models with *o*_*p*_(1) remainders.

2 The *p*-values are not always calibrated in practice, because the test assumes that *α*_*j*_ is fixed. We do not use this test for formal inference, but only for visualizing and summarizing the degree of Mendelianism.

## Notes

### Competing Interest Statement

The authors have declared no competing interest.

### Author Declarations

The study used only openly available human data that were originally located at: Pan-UK Biobank (https://pan.ukbb.broadinstitute.org/)

### Summary of Updates

Refined theorems; improved explanation on coefficient interpretation; added simulation results with latent confounding

